# Large language model consensus for reliable research cohort construction from radiology reports: a retrospective cohort study

**DOI:** 10.64898/2026.10.06.26364312

**Authors:** Wushuang Rui, Jiajian Ma, Valentin Stepanov, Arianna Bunnell, Michael Lan, Jenny Chen, Roshni R. Patel, Dmitry S. Novikov, Els Fieremans, Yiqiu Shen

## Abstract

**Background:** Large-scale retrospective studies often require researchers to adjudicate outcomes or cohort eligibility from unstructured clinical records. Manual review can provide reliable labels but is difficult to scale. We developed and evaluated a consensus workflow using multiple large language models (LLMs) to automatically assign high-confidence outcome labels while deferring ambiguous cases for manual review.

**Methods:** We included 6,718 brain MRI reports from 5,856 patients. The expert-adjudicated reference set included 680 reports from 675 patients: 372 normal (age-appropriate without significant intracranial pathology), 90 white matter hyperintensity-only (WMH-only; isolated nonspecific WMHs potentially ambiguous for binary classification), and 218 abnormal (other intracranial pathology) reports. We evaluated proprietary-inclusive (Qwen3/Kimi-Linear/GPT-5) and open-source (Qwen3/Kimi-Linear/DeepSeek-R1) ensembles against individual LLMs. Ensembles assigned a normal or abnormal label only when all models agreed; all other reports were classified as uncertain and deferred for manual review. Individual LLMs assigned normal or abnormal predictions and classified WMH-only predictions as uncertain. In the full cohort, we compared the prevalence of active central nervous system (CNS)-related International Classification of Diseases, Tenth Revision (ICD-10) diagnoses across the normal, uncertain, and abnormal groups generated by each ensemble.

**Results:** In the expert-adjudicated reference set, both ensembles achieved 100% precision for automatically assigned normal and abnormal reports. GPT-5 was the strongest individual LLM but had lower precision than either ensemble, with normal and abnormal precision of 98.9% and 94.1%, respectively, and 17 expert-label mismatches versus none for the consensus ensembles. In the full cohort, 1.6% of reports classified as normal were associated with CNS-related ICD-10 diagnoses, compared with 29.7% of uncertain and 51.3% of abnormal reports for the proprietary-inclusive ensemble; corresponding rates were 1.6%, 31.1%, and 51.6% for the open-source ensemble.

**Conclusions:** Requiring unanimous agreement across multiple LLMs enabled high-precision automatic adjudication while directing ambiguous cases to manual review. This strategy may reduce the burden of manual adjudication and support scalable construction of high-purity research cohorts from unstructured clinical text.

## Background

Large neuroimaging studies have traditionally relied on purpose-built research imaging cohorts, such as UK Biobank, the Alzheimer’s Disease Neuroimaging Initiative, Human Connectome Project, and the Adolescent Brain Cognitive Development Study, in which participants are recruited and characterized according to predefined study protocols [1–4]. Routine clinical imaging archives represent a complementary and substantially underused source of brain magnetic resonance imaging (MRI) data [5, 6]. These archives contain large numbers of examinations acquired across diverse clinical populations and may support downstream case-control studies, normative modeling, and brain-age modeling [5–7]. However, because these examinations were acquired for clinical care rather than research, they are not routinely labeled as normal or abnormal for downstream use [6, 8].

Accurate identification of radiographically normal and abnormal examinations is therefore an important step toward constructing research datasets from clinical MRI archives. Normal examinations are characterized by age-appropriate brain appearance without clinically significant intracranial abnormalities, whereas abnormal examinations contain clear pathological intracranial findings [9–11]. Misclassification of abnormal examinations as normal can compromise dataset validity and bias downstream analyses [12–17]. Although expert review can make this distinction, reviewing large numbers of examinations is labor-intensive and difficult to scale across health-system imaging archives [18–21]. Automated classification could therefore support efficient and reproducible identification of radiographically normal and abnormal examinations at scale [19, 20, 22–24].

Scalable identification of normal and abnormal examinations from clinical imaging archives remains challenging because routinely available clinical information does not reliably provide examination-level labels for brain MRI findings. International Classification of Diseases, Tenth Revision (ICD-10) diagnosis codes are widely available but indirect; they may reflect the patient’s symptoms, medical history, or previously recorded diagnoses rather than the imaging findings of the corresponding brain MRI examination [18, 25–27]. Compared with ICD-10 diagnosis codes, brain MRI radiology reports provide a more direct description of radiological findings [19, 20, 28, 29]. However, brain MRI reports are written as free text and vary in wording, making it difficult to classify large numbers of reports consistently with traditional rule-based or bag-of-words approaches [19, 28, 30, 31]. Reliable classification of clinical MRI examinations requires scalable methods for deriving examination-level labels directly from brain MRI reports.

A further challenge is that the definition of a normal brain MRI depends on the research question. For example, mild white matter hyperintensities (WMHs) or age-related brain atrophy may be acceptable in brain-age studies when age-appropriate, whereas strictly normal cohorts used to define normative brain structure may exclude these findings [12, 14, 32]. Even in brain-age studies that allow age-related changes, isolated nonspecific WMHs can be difficult to classify. Whether these WMHs are considered normal may depend on patient age, lesion burden, and anatomical distribution [33–44]. Classifying these ambiguous examinations as normal may introduce potentially pathological findings into the normal MRI group, whereas classifying them as abnormal may group mild age-related changes with clearly pathological examinations [33, 35, 44]. Reliable use of clinical MRI data therefore requires separating ambiguous examinations from those that are clearly radiographically normal or abnormal.

Recent advances in natural language processing (NLP) and large language models (LLMs) have made it possible to classify large numbers of brain MRI examinations from their radiology reports [18–20, 29, 45–47]. A key challenge, however, is that reports with ambiguous findings may not have a universally applicable classification, so their labels may vary by study objective and expert reviewer [48–52]. Our hypothesis in this study was that different LLMs disagree more often on ambiguous cases and that model disagreement can identify reports requiring manual review. Based on this hypothesis, we developed a multi-LLM consensus workflow that assigns a normal or abnormal label only when all models agree; all other reports are classified as uncertain and deferred for manual review. We evaluated whether this selective approach could support accurate, semi-automated identification of normal and abnormal brain MRI from radiology reports.

## Methods

### Cohort description

We conducted a retrospective cohort study to develop and evaluate individual-and multi-LLM strategies to determine if a brain MRI report qualifies as a control (“normal”) or not (“abnormal”). All brain MRI reports included in this study were collected from NYU Langone Health, an academic tertiary-care center in New York. The Institutional Review Board of NYU Grossman School of Medicine approved the study (IRB i14-01224 and i25-01143) and waived the requirement for informed consent. Reports and linked diagnosis-code data were de-identified in accordance with institutional policies and HIPAA requirements.

The cohort included consecutive reports from brain MRI examinations performed at an NYU Langone Health outpatient imaging center between October 2014 and October 2020. These reports were derived from examinations performed using a routine brain MRI protocol. Reports were excluded if they did not describe a brain MRI examination, lacked an impression section, or contained fewer than 25 words (Additional file 1: Fig. S1). After application of these exclusion criteria, the final study cohort comprised 6,718 brain MRI reports. Patient age, recorded sex, and linked central nervous system (CNS)-related ICD-10 diagnoses were obtained from the institutional electronic health record (EHR). For cohort characterization, CNS diagnoses were grouped into seven broad categories: vascular, traumatic, neoplastic, infectious/inflammatory/immune-mediated, congenital/developmental, cerebrospinal fluid circulation/structural, and neurodegenerative. Definitions and the ICD-10 codes included in each category are provided in Additional file 1: Table S1.

### Report preprocessing

Report preprocessing comprised two steps: (1) section segmentation using regular expressions and (2) report conversion to JavaScript Object Notation (JSON) format using an LLM (Fig. 1A). Custom regular-expression templates were applied to parse each radiology report into five standardized sections: clinical indication, technique, findings, impression, and other. For each evaluated LLM, the parsed report text was converted into JSON by the same model that subsequently performed classification, using a fixed prompt provided verbatim in Additional file 1: Methods and a standardized JSON schema. The JSON schema captured clinical indication, report-described lesion category (e.g., hemorrhage or arteriovenous malformation), anatomical location, lesion size, imaging characteristics, and incidental findings. If not clearly stated in the report, the corresponding field was encoded as null rather than inferred.

**Fig. 1.**
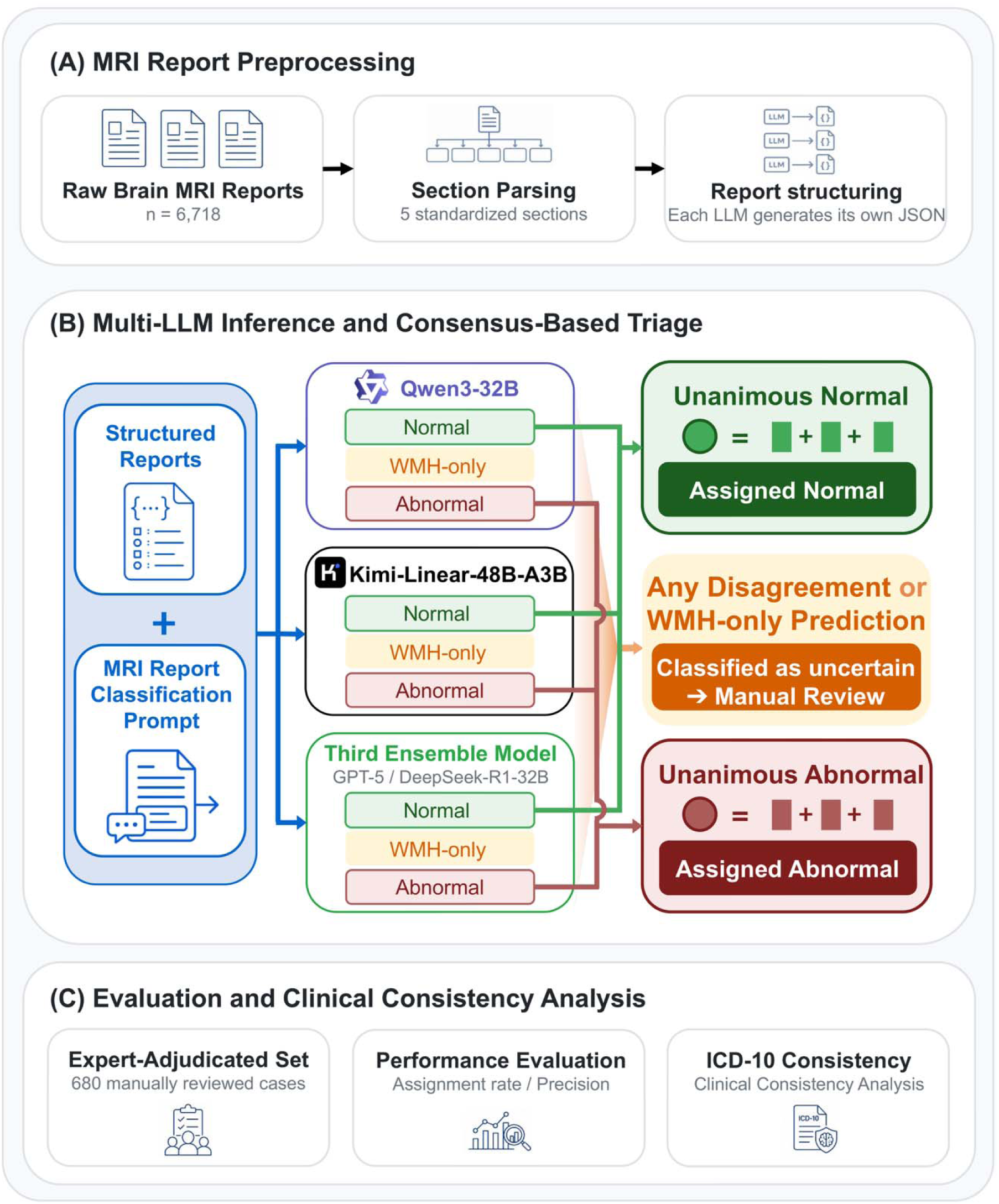
Multi-LLM consensus workflow for selective brain MRI report classification. (A) Brain MRI reports were parsed into standardized sections, and each LLM independently generated a structured representation using the same predefined JSON schema and prompt. (B) Three separately deployed LLMs classified each report as normal, WMH-only, or abnormal. Reports were assigned only when all models unanimously predicted normal or abnormal; reports with any WMH-only prediction or model disagreement were classified as uncertain and deferred for manual review. (C) The workflow was evaluated using an expert-adjudicated reference set, assigned-label performance metrics, and within-institution consistency analysis using CNS-related ICD-10 diagnoses.

### Expert reference standard

From the full study cohort of 6,718 reports, we assembled a reference set of 680 brain MRI reports for model evaluation. The reference set included 380 reports from an earlier internal project and 300 additional reports selected from the remaining study cohort to capture a broader range of brain MRI abnormalities. The 380 reports had been labeled as normal by a medical student (M.P.) in an earlier internal project and subsequently reviewed with a radiologist (T.S.). To select the additional 300 reports, we used lesion categories derived from DeepSeek-R1-32B–generated JSON representations of the full study cohort, as described above. We ranked these lesion categories by frequency and selected one previously unselected report representing each of the 300 most frequent categories (Additional file 1: Table S2). No reports in the reference set were used to train or fine-tune the evaluated LLMs. The reference set was used to evaluate classification reliability rather than to estimate workflow coverage in the source clinical population.

One board-certified neuroradiologist (R.R.P.), one preliminary-year diagnostic radiology resident (M.L.), and one postdoctoral fellow with 9 years of neuroimaging experience (V.S.) initially reviewed each report independently and assigned one of three labels: normal, white matter hyperintensity–only (WMH-only), or abnormal. Discrepant or borderline cases were subsequently reviewed jointly to establish the final consensus label. Reports were labeled normal if they showed an age-appropriate brain appearance without clinically significant intracranial parenchymal abnormalities. Reports were labeled WMH-only if they showed isolated nonspecific WMHs without other clinically significant intracranial abnormalities. Reports were labeled abnormal if they showed pathological intracranial findings beyond isolated nonspecific WMHs. Reviewers were blinded to all LLM outputs and ICD-10 codes throughout independent review and consensus adjudication.

### Prompt design

All models used the same fixed zero-shot classification prompt (Listing 1). The prompt consisted of five components: (1) a role instruction defining the model as a clinical neuroimaging specialist with expertise in neuroradiology, neurology, and neurosurgery; (2) a task specification that required examination-level classification based on patient age at MRI and the structured report representation generated during preprocessing; (3) operational definitions of normal, WMH-only, and abnormal examinations to standardize classification boundaries across models; (4) decision and exclusion rules that required the models to consider all available information, identify clinically significant intracranial abnormalities, and avoid classifying benign incidental findings, extracranial findings, or anatomical variants as abnormal; and (5) a constrained output format that required a single numeric label corresponding to normal, WMH-only, or abnormal.

**Listing 1. Fixed zero-shot prompt used for examination-level classification of brain MRI radiology reports**

ROLE

You are a certified clinical neuroimaging specialist with multidisciplinary expertise, including:

• A neuroradiologist with 15+ years of experience interpreting brain MRIs, skilled in detecting subtle and clinically significant intracranial abnormalities;

• A neurologist specializing in multiple sclerosis, neurodegenerative diseases, and conditions with brain atrophy; and

• A neurosurgeon focusing on functional neurosurgery, including Gamma Knife, high-intensity focused ultrasound, and deep brain stimulation.

You integrate imaging findings with clinical context using a strictly evidence-based approach.

TASK

Analyze a JSON-formatted brain MRI radiology report and classify the examination based only on clinically significant intracranial findings. Follow all definitions and exclusion rules strictly.

CLASSIFICATION CATEGORIES

Output must be one of: 1, 2, or 3.

1 — Normal MRI

• No abnormal intracranial parenchymal findings.

• Brain structures are age-appropriate and within normal limits.

2 — White matter hyperintensity–only

• Nonspecific white matter lesions; or

• Chronic ischemic changes without restricted diffusion or stroke sequelae, such as encephalomalacia, gliosis, or cavitation.

• No other abnormalities are present.

3 — Abnormal MRI

• Any intracranial parenchymal abnormality other than, or in addition to, nonspecific white matter lesions, including infarction, restricted diffusion, stroke sequelae, mass, hemorrhage, hydrocephalus, or Chiari malformation.

• A history of brain surgery or intracranial treatment, including deep brain stimulation or Gamma Knife treatment, unless clearly unrelated to brain pathology.

DECISION RULES

1. Focus on brain parenchymal findings. Ignore findings involving non-brain parenchymal structures, including intracranial blood vessels, sinuses, mastoids, paranasal regions, and the spine. However, include intracranial tumors that are not parenchymal, including meningiomas, cranial nerve schwannomas, and pituitary tumors.

2. The following findings should not independently trigger Category 3:

- Small chronic microbleeds or susceptibility foci in elderly patients, unless they are numerous, lobar, traumatic in a younger patient, or associated with parenchymal changes such as contusion or encephalomalacia.
- Findings explicitly described as benign, incidental, or clinically insignificant. Findings likely to be benign should not be classified as Category 3 solely because they are not included in the exclusion list.

3. Consider all sections of the report equally. Do not prioritize the Impression or Conclusion. Base the classification on the complete report.

EXCLUSION LIST: FINDINGS THAT MAY REMAIN NORMAL

A. Benign intracranial findings

• Small or asymptomatic pineal cyst

• Arachnoid, retention, or other small cyst

• Developmental venous anomaly

• Capillary telangiectasia

• Dilated perivascular spaces

• Perineural cyst

• Asymmetric fatty marrow

• Benign intraosseous hemangioma

• Incidental cyst smaller than 1 cm

• Sulcal or ventricular enlargement explicitly described as age-related

• Mild volume loss

B. Extracranial findings, when unrelated to an intracranial abnormality

• Sinus or mastoid mucosal thickening or cysts

• Skull-base findings involving structures such as the jugular foramen or internal auditory canal

• Vertebral or disc findings from C1 through T1

• Findings involving the parotid gland, nasopharynx, pterygoid region, or fossa of Rosenmüller

C. Nonpathological anatomical variants

• Hypoplastic transverse or sigmoid sinus

• Small vertebral artery

• Mild dolichoectasia

• Sinus air-fluid levels

• Mucus retention cysts

INPUT

PATIENT AGE: [age at MRI]; MRI REPORT: [JSON-formatted brain MRI radiology report]

REQUIRED OUTPUT

Return only one numeric label: 1, 2, or 3.

### Classification models and inference

We defined both proprietary-inclusive and open-source LLM ensembles for consensus classification experiments. Each ensemble consisted of three distinct LLMs. The proprietary-inclusive ensemble included Qwen3-32B, Kimi-Linear-48B-A3B, and GPT-5 [53–55]. The open-source ensemble replaced GPT-5 with DeepSeek-R1-Distill-Qwen-32B (DeepSeek-R1-32B) [56]. Local inference used vLLM, Transformers, and PyTorch [57–59]. Qwen3-32B, Kimi-Linear-48B-A3B, and DeepSeek-R1-32B were run without quantization in Brain floating-point 16-bit format (bfloat16) precision. Qwen3-32B and DeepSeek-R1-32B were run using Python 3.10.16, vLLM 0.9.0, Transformers 4.52.3, and PyTorch 2.7.0+cu118 on one NVIDIA A100 GPU, whereas Kimi-Linear-48B-A3B was run using Python 3.10.21, vLLM 0.11.2, Transformers 4.57.6, and PyTorch 2.9.0+cu128 with tensor parallelism across four NVIDIA A100 80-GB GPUs. For all three locally deployed models, the random seed was set to 42. GPT-5 was accessed on September 5, 2026, through the institutional endpoint gpt-5/v1.0.0 using OpenAI Python 2.30.0 and Python 3.10.20; the provider-specific underlying snapshot was not exposed, and no random seed was explicitly set. For all models, temperature was set to 0.2, top_p to 1.0, and the maximum output length to 512 tokens. Each report contributed one final classification from each model.

### Classification strategies

#### Individual-model baselines

Each of the four LLMs—Kimi-Linear-48B-A3B, Qwen3-32B, GPT-5, and DeepSeek-R1-32B—was also evaluated as an individual-model baseline. Reports predicted as normal or abnormal were assigned the corresponding label, whereas reports predicted as WMH-only were classified as uncertain and deferred for manual review.

#### Majority-voting comparator

Majority voting was evaluated as a comparator to the multi-LLM consensus workflow for each LLM ensemble. Under majority voting, any WMH-only prediction resulted in classification as uncertain; otherwise, the report was assigned the normal or abnormal label supported by the majority of models. This comparison was used to isolate the effect of requiring unanimous rather than majority agreement.

#### Multi-LLM consensus workflow

Multi-LLM consensus was the primary approach for examination-level classification in each LLM ensemble (Fig. 1B). Reports were assigned as normal or abnormal only when all three models agreed on the same normal or abnormal label. Any WMH-only prediction or model disagreement resulted in classification as uncertain and was deferred for manual review.

### ICD-10–based analysis

#### Definition of ICD-10 status

To define CNS-related ICD-10 status for each report, we matched ICD-10 codes in the EHR to a curated list of CNS diagnoses considered relevant to abnormal brain MRI findings. The list of relevant diagnoses was assembled by a postdoctoral fellow with 9 years of neuroimaging experience (V.S.) (Additional file 1: Table S3). A code was counted only if it was recorded within 10 days before or after the MRI examination and listed as an active diagnosis for the encounter linked to that examination. Codes marked as historical, possible, suspected, or ruled out were excluded. An examination was then classified as ICD-10-positive if at least one qualifying active CNS-related diagnosis was present and ICD-10-negative otherwise.

#### ICD-10–based comparator

Within the expert-adjudicated reference set, ICD-10-positive examinations were classified as abnormal and ICD-10-negative examinations as normal. This provided a binary ICD-10–based classification strategy for comparison with the LLM methods.

#### Full-cohort comparison with CNS-related ICD-10 diagnoses

Across the full cohort, we calculated the prevalence of active CNS-related ICD-10 diagnoses among examinations in the normal, uncertain, and abnormal groups produced by each multi-LLM consensus ensemble. We examined whether this prevalence increased from the normal group to the uncertain group and was highest in the abnormal group.

### Statistical analysis

The brain MRI examination was the unit of analysis, and each examination was represented by one brain MRI report. The individual LLM baselines, majority-voting comparator, multi-LLM consensus workflow, and ICD-10-based comparator were evaluated against the examination-level labels in the 680-report expert-adjudicated reference set. Because the LLM-based approaches allowed reports to be classified as uncertain rather than forcing every report into a normal or abnormal category, we evaluated the accuracy of automatically assigned reports and the percentage of reports assigned without manual review. We also calculated the proportion of expert-labeled WMH-only examinations assigned to the uncertain group. The ICD-10 comparator had no uncertain category and therefore classified every report as either normal or abnormal.

The assignment rate was the percentage of reports assigned a normal or abnormal label rather than classified as uncertain. Among automatically assigned reports, assignment accuracy and assignment error rate were the proportions of assignments that matched and did not match the expert-adjudicated label, respectively. Normal and abnormal precision were the proportions of normal and abnormal assignments, respectively, that matched the corresponding expert label. Normal and abnormal recall were defined as the proportions of expert-normal and expert-abnormal examinations correctly assigned as normal and abnormal, respectively, without manual review. The number of assignment errors was defined as the total number of incorrect assignments; expert WMH-only reports assigned as either normal or abnormal were therefore treated as errors. The WMH-only deferral rate was the proportion of expert WMH-only reports classified as uncertain. For the ICD-10 comparator, the assignment rate was 100% and the WMH-only deferral rate was 0% by design. We computed 95% bootstrap confidence intervals using 2,000 patient-level bootstrap replicates.

To characterize prediction patterns within the uncertain group, we separated uncertain reports into those with unanimous WMH-only predictions and those with model disagreement. For each pattern, we examined the distribution of expert-adjudicated normal, WMH-only, and abnormal labels. We also calculated, among expert WMH-only reports, the proportions identified by unanimous WMH-only prediction and by model disagreement.

## Results

### Cohort characteristics

After exclusions, the cohort contained 6,718 brain MRI reports from 5,856 unique patients: 680 expert-adjudicated reference reports and 6,038 non-reference reports (Table 1). The expert-adjudicated reference set contained 372 normal, 90 WMH-only, and 218 abnormal reports. Among the 5,856 patients, 694 (11.9%) contributed two or more reports, with a maximum of 6 reports per patient. The cohort included patients across a broad age range and both recorded sexes, with varied MRI scanner use and approximately one-third of examinations having an active CNS-related ICD-10 diagnosis.

**Table 1.** Cohort characteristics and active CNS ICD-10 diagnostic categories.

|  | Full cohort | Non-reference set | Reference set |
| --- | --- | --- | --- |
| **Patients, n** | 5856 | 5226 | 675 |
| **MRI examinations, n** | 6718 | 6038 | 680 |
| **Patients with $\geq 2$ MRI examinations, n (%)** | 694 (11.9%) | 654 (12.5%) | 5 (0.7%) |
| **MRI scanner (scan-level), n (%)** |  |  |  |
| MAGNETOM Aera | 90 (1.3%) | 89 (1.5%) | 1 (0.1%) |
| Biograph mMR | 1 (0.0%) | 0 (0.0%) | 1 (0.1%) |
| MAGNETOM Prisma fit | 2137 (31.8%) | 1877 (31.1%) | 260 (38.2%) |
| MAGNETOM Skyra | 4490 (66.8%) | 4072 (67.4%) | 418 (61.5%) |
| **Sex** |  |  |  |
| Female | 3450 (58.9%) | 3042 (58.2%) | 438 (64.9%) |
| Male | 2406 (41.1%) | 2184 (41.8%) | 237 (35.1%) |
| **Age, years** | 50.61 ± 20.55 | 50.95 ± 20.80 | 47.59 ± 17.95 |
| **Active CNS ICD-10 Diagnoses** |  |  |  |
| Yes | 2223 (33.1%) | 2105 (34.9%) | 118 (17.4%) |
| No | 4495 (66.9%) | 3933 (65.1%) | 562 (82.6%) |
| **Active CNS ICD-10 Categories** |  |  |  |
| Vascular | 906 (13.5%) | 826 (13.7%) | 80 (11.8%) |
| Traumatic | 245 (3.6%) | 213 (3.5%) | 32 (4.7%) |
| Neoplastic | 403 (6.0%) | 378 (6.3%) | 25 (3.7%) |
| Infectious/Inflammatory/Immune-mediated | 1441 (21.4%) | 1380 (22.9%) | 61 (9.0%) |
| Congenital/Developmental | 49 (0.7%) | 49 (0.8%) | 0 (0.0%) |
| CSF circulation/Structural | 522 (7.8%) | 482 (8.0%) | 40 (5.9%) |
| Neurodegenerative | 211 (3.1%) | 190 (3.1%) | 21 (3.1%) |

Values are counts unless otherwise specified; age is reported as mean ± standard deviation. Sex was summarized at the patient level, whereas age, scanner characteristics, and ICD-10 diagnoses were summarized at the examination level. Some patients contributed to both subsets, so patient counts do not sum to the full-cohort total. Diagnosis categories were not mutually exclusive; an examination could contribute to more than one category. CSF, cerebrospinal fluid.

### Performance of the multi-LLM consensus workflow

Assignment rates were 70.7% and 67.4% on the reference set and 52.0% and 48.6% across the full cohort for the proprietary-inclusive and open-source ensembles, respectively. On the reference set, both ensembles achieved 100.0% assignment accuracy and 100.0% precision for both normal and abnormal classifications (0/481 and 0/458 incorrect assignments) (Table 2). Normal recall was 85.8% (95% confidence interval (CI), 82.2%–89.1%) and 86.3% (95% CI, 82.8%–89.6%), whereas abnormal recall was 74.3% (95% CI, 68.6%–80.0%) and 62.8% (95% CI, 56.3%–69.3%) for the proprietary-inclusive and open-source ensembles, respectively (Additional file 1: Table S4). Both ensembles deferred all 90 expert-adjudicated WMH-only reports. In comparison, the ICD-10–based comparator assigned all reports but had substantially lower assignment accuracy (70.7%; 95% CI, 67.2%–74.0%) and normal precision (65.8%; 95% CI, 61.8%–69.7%), with no WMH-only deferral.

**Table 2.**
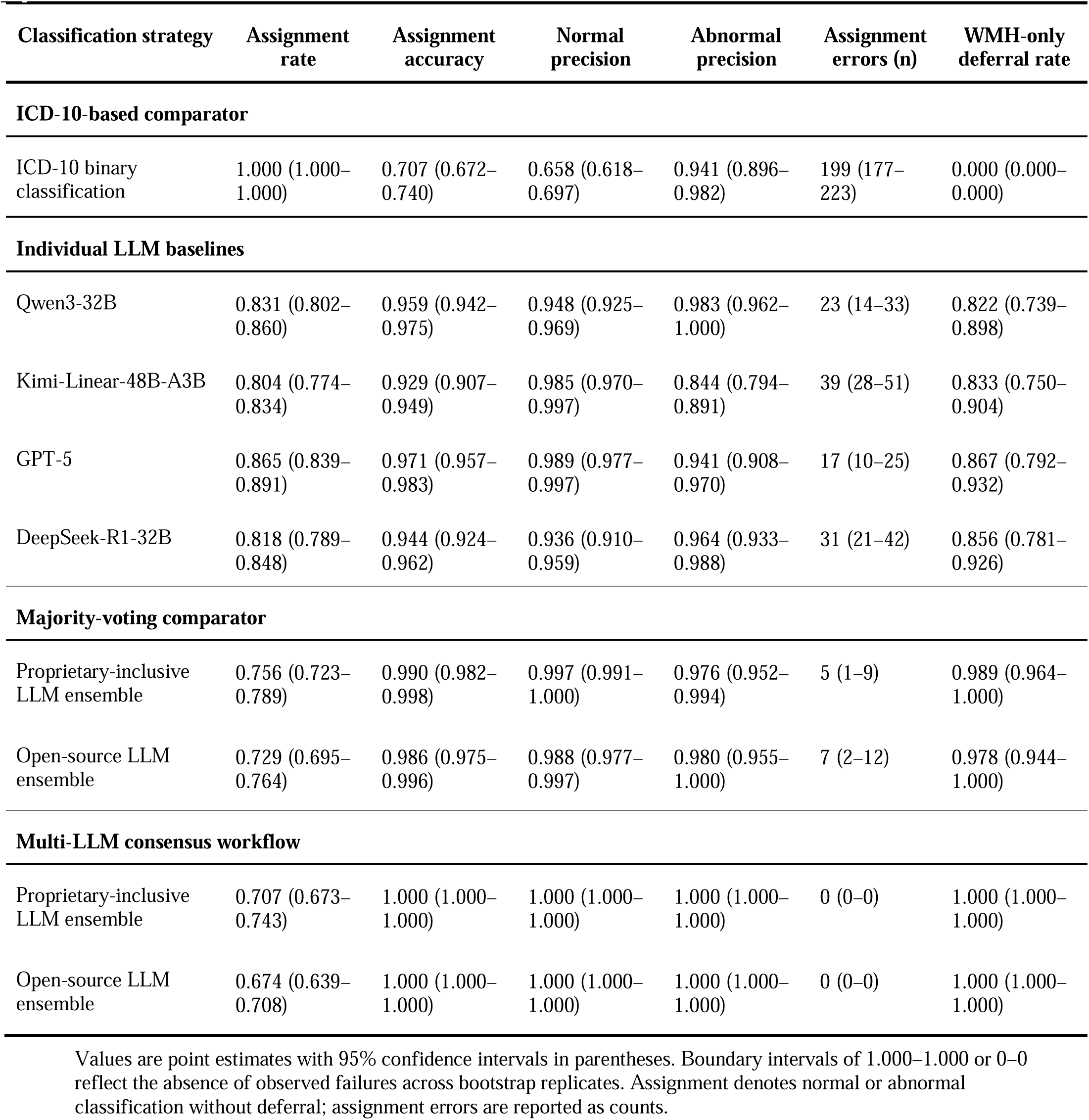
Reference-set performance of ICD-10 comparator, individual LLMs, majority-voting ensembles, and multi-LLM consensus ensembles.

| Classification strategy | Assignment rate | Assignment accuracy | Normal precision | Abnormal precision | Assignment errors (n) | WMH-only deferral rate |
| --- | --- | --- | --- | --- | --- | --- |
| **ICD-10-based comparator** |  |  |  |  |  |  |
| ICD-10 binary classification | 1.000 (1.000–1.000) | 0.707 (0.672–0.740) | 0.658 (0.618–0.697) | 0.941 (0.896–0.982) | 199 (177–223) | 0.000 (0.000–0.000) |
| **Individual LLM baselines** |  |  |  |  |  |  |
| Qwen3-32B | 0.831 (0.802–0.860) | 0.959 (0.942–0.975) | 0.948 (0.925–0.969) | 0.983 (0.962–1.000) | 23 (14–33) | 0.822 (0.739–0.898) |
| Kimi-Linear-48B-A3B | 0.804 (0.774–0.834) | 0.929 (0.907–0.949) | 0.985 (0.970–0.997) | 0.844 (0.794–0.891) | 39 (28–51) | 0.833 (0.750–0.904) |
| GPT-5 | 0.865 (0.839–0.891) | 0.971 (0.957–0.983) | 0.989 (0.977–0.997) | 0.941 (0.908–0.970) | 17 (10–25) | 0.867 (0.792–0.932) |
| DeepSeek-R1-32B | 0.818 (0.789–0.848) | 0.944 (0.924–0.962) | 0.936 (0.910–0.959) | 0.964 (0.933–0.988) | 31 (21–42) | 0.856 (0.781–0.926) |
| **Majority-voting comparator** |  |  |  |  |  |  |
| Proprietary-inclusive LLM ensemble | 0.756 (0.723–0.789) | 0.990 (0.982–0.998) | 0.997 (0.991–1.000) | 0.976 (0.952–0.994) | 5 (1–9) | 0.989 (0.964–1.000) |
| Open-source LLM ensemble | 0.729 (0.695–0.764) | 0.986 (0.975–0.996) | 0.988 (0.977–0.997) | 0.980 (0.955–1.000) | 7 (2–12) | 0.978 (0.944–1.000) |
| **Multi-LLM consensus workflow** |  |  |  |  |  |  |
| Proprietary-inclusive LLM ensemble | 0.707 (0.673–0.743) | 1.000 (1.000–1.000) | 1.000 (1.000–1.000) | 1.000 (1.000–1.000) | 0 (0–0) | 1.000 (1.000–1.000) |
| Open-source LLM ensemble | 0.674 (0.639–0.708) | 1.000 (1.000–1.000) | 1.000 (1.000–1.000) | 1.000 (1.000–1.000) | 0 (0–0) | 1.000 (1.000–1.000) |

Values are point estimates with 95% confidence intervals in parentheses. Boundary intervals of 1.000–1.000 or 0–0 reflect the absence of observed failures across bootstrap replicates. Assignment denotes normal or abnormal classification without deferral; assignment errors are reported as counts.

### Composition of assigned and deferred reports

All assigned normal and abnormal reports matched the corresponding expert labels in both ensembles (Fig. 2). In the proprietary-inclusive ensemble, the uncertain group comprised 53 expert-normal reports (26.6%), 90 WMH-only reports (45.2%), and 56 expert-abnormal reports (28.1%). In the open-source ensemble, the corresponding composition was 51 expert-normal reports (23.0%), 90 WMH-only reports (40.5%), and 81 expert-abnormal reports (36.5%). Thus, the uncertain group comprised a mixture of expert-normal, WMH-only, and expert-abnormal reports, including all WMH-only cases.

**Fig. 2.**
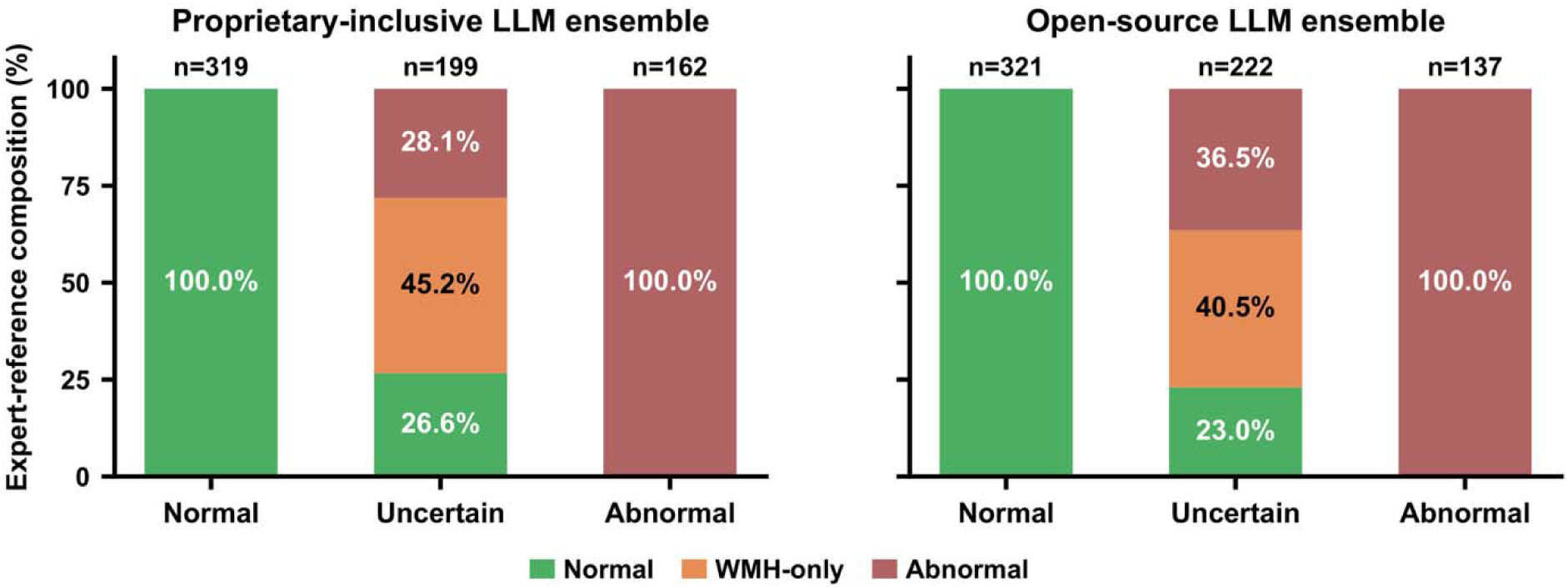
Expert-adjudicated label composition across multi-LLM consensus output categories. Stacked bars show the expert-adjudicated label composition of reports assigned as normal, deferred as uncertain, or assigned as abnormal by the proprietary-inclusive (left) and open-source (right) ensembles. Colors denote normal, WMH-only, and abnormal expert-reference labels. Numbers above the bars indicate the number of reports in each output category, and percentages within the bars indicate their expert-reference composition.

### Prediction patterns in the uncertain group

Within the uncertain group, model disagreement was more common than unanimous WMH-only predictions in both ensembles (131 vs 68 reports in the proprietary-inclusive ensemble and 149 vs 73 in the open-source ensemble; Fig. 3). Unanimous WMH-only predictions corresponded to expert WMH-only labels in 91.2% and 82.2% of cases, respectively, although this precision was lower than that of unanimous normal and abnormal assignments (100.0%). In contrast, model-disagreement cases showed a more diverse expert-label composition: in the proprietary-inclusive ensemble, 38.9% were expert-normal, 21.4% expert WMH-only, and 39.7% expert-abnormal; in the open-source ensemble, the corresponding proportions were 32.9%, 20.1%, and 47.0%. Among expert WMH-only cases, 68.9% and 66.7% were identified by unanimous WMH-only predictions, with an additional 31.1% and 33.3% captured through model disagreement. Together, the two deferral signals captured different case compositions: WMH-only predictions primarily identified prespecified WMH-only cases, whereas model disagreement captured the remaining WMH-only cases together with a broader mixture of expert-normal and expert-abnormal reports.

**Fig. 3.**
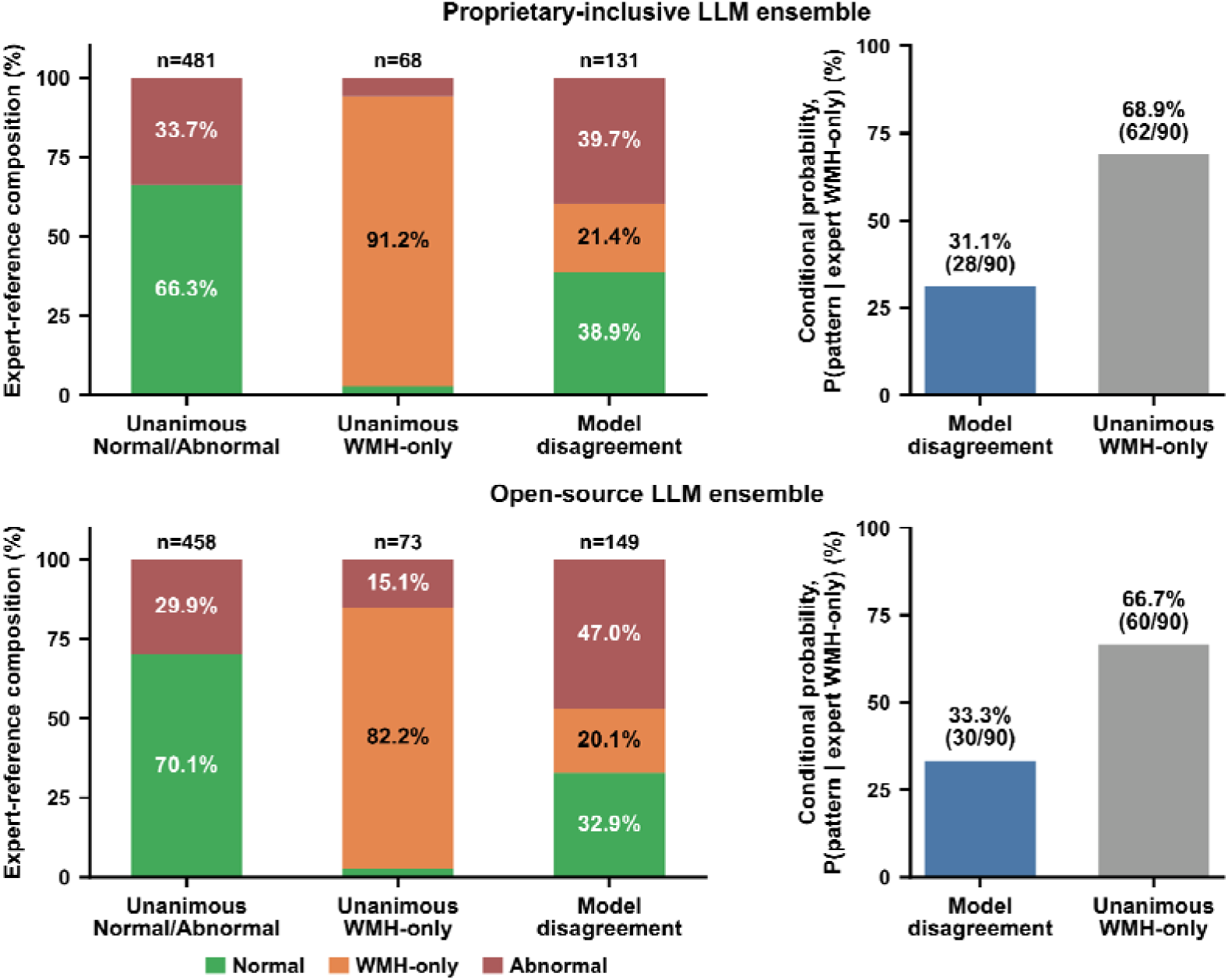
Expert-adjudicated label composition across multi-LLM prediction patterns. For the proprietary-inclusive (top) and open-source (bottom) large language model (LLM) ensembles, left-hand stacked bars show the expert-adjudicated label composition of reports with unanimous normal or abnormal predictions, unanimous white matter hyperintensity-only (WMH-only) predictions, or model disagreement. Colors denote normal, WMH-only, and abnormal expert-reference labels. Numbers above the bars indicate the number of reports in each prediction-pattern category, and percentages within the bars indicate their expert-reference composition. Right-hand bars show, among the 90 expert WMH-only reports, the proportions resulting in model disagreement or unanimous WMH-only prediction; counts are shown in parentheses.

### Comparison with individual LLM predictions

Individual LLM baselines achieved assignment accuracies of 92.9%–97.1% and produced 17–39 incorrect assignments (Table 2). In contrast, both multi-LLM consensus ensembles achieved 100.0% assignment accuracy and zero incorrect assignments, with 100.0% precision for both normal and abnormal classifications and 100.0% WMH-only deferral, albeit at lower assignment rates. Compared with the best-performing individual model in each ensemble, multi-LLM consensus intercepted all 17 GPT-5 errors in the proprietary-inclusive ensemble and all 23 Qwen3-32B errors in the open-source ensemble (100.0% for both; Fig. 4). Most intercepted errors were expert WMH-only reports, indicating that consensus deferral preferentially captured ambiguous, error-prone cases.

**Fig. 4.**
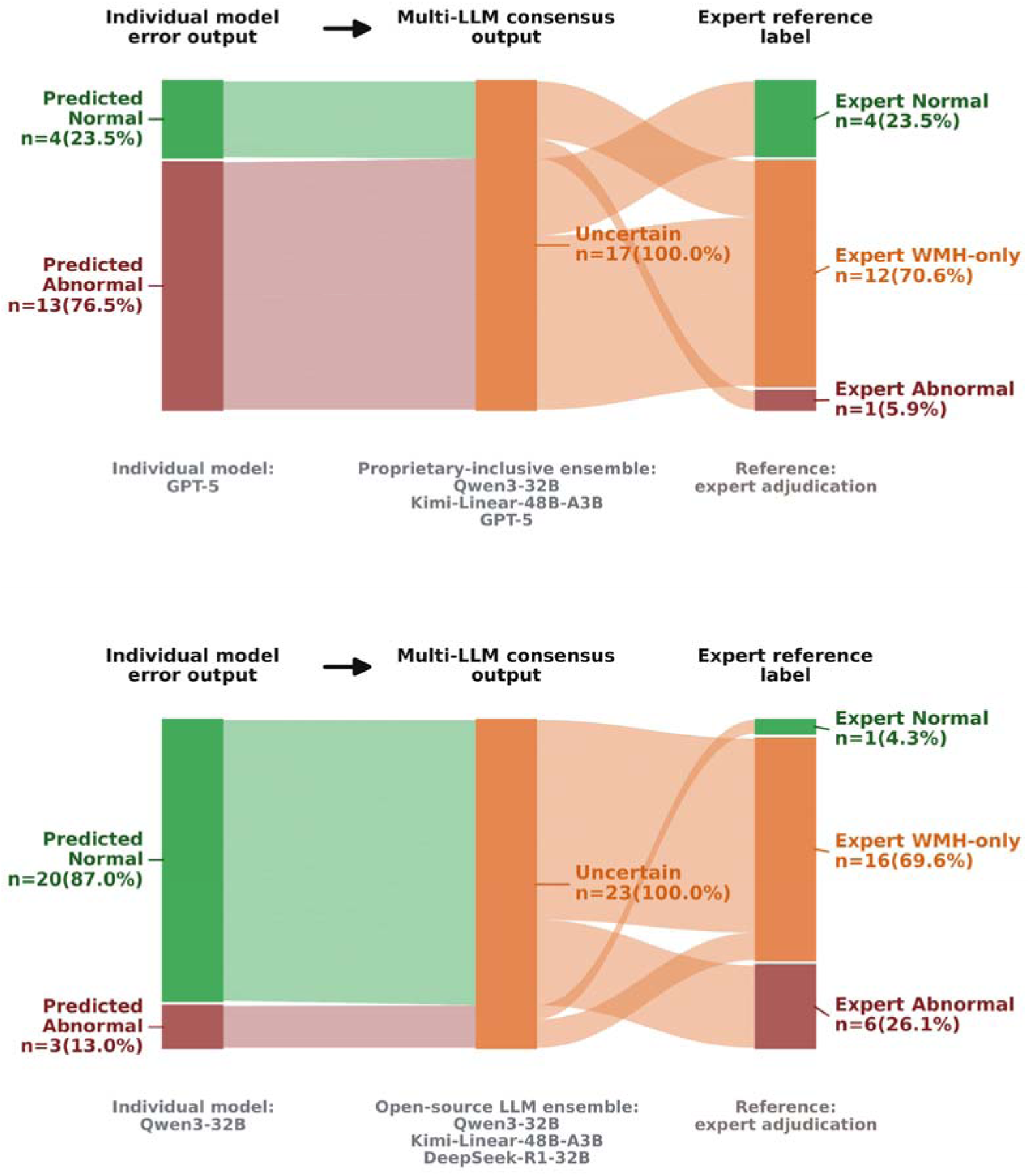
Deferral of single-model assignment errors by multi-LLM consensus. Alluvial diagrams trace incorrectly assigned individual model predictions through the corresponding multi-LLM consensus workflow and expert-adjudicated reference labels. The top panel shows GPT-5 errors evaluated by the proprietary-inclusive ensemble comprising Qwen3-32B, Kimi-Linear-48B-A3B, and GPT-5. The bottom panel shows Qwen3-32B errors evaluated by the open-source ensemble comprising Qwen3-32B, Kimi-Linear-48B-A3B, and DeepSeek-R1-32B. The consensus workflow deferred all single-model errors as uncertain (17/17 for GPT-5 and 23/23 for Qwen3-32B). Most involved expert-adjudicated white matter hyperintensity-only (WMH-only) reports (12/17 [70.6%] and 16/23 [69.6%], respectively).

### Assignment coverage and error risk across selective strategies

Figure 5 compares the coverage–risk tradeoff across the individual LLM baseline, majority-voting comparator, and multi-LLM consensus workflow and examines the error rate among reports newly assigned as the agreement criteria are progressively relaxed. In the proprietary-inclusive ensemble, the assignment rate decreased from 86.5% for GPT-5 to 75.6% for majority voting and 70.7% for multi-LLM consensus, while the corresponding assignment error rate decreased from 2.9% to 1.0% and 0.0%, respectively. In the open-source ensemble, the assignment rate decreased from 83.1% for Qwen3-32B to 72.9% for majority voting and 67.4% for multi-LLM consensus, while the error rate decreased from 4.1% to 1.4% and 0.0%, respectively (Fig. 5A). Reports retained under multi-LLM consensus had no errors in either ensemble (0/481 and 0/458). Among reports newly assigned by majority voting relative to consensus, majority-vote error rates were 15.2% (5/33) and 18.4% (7/38) for the proprietary-inclusive and open-source ensembles, respectively. Among reports additionally assigned only by the corresponding individual-model baseline, individual-model error rates were 14.9% (11/74) and 24.6% (17/69), respectively (Fig. 5B). Thus, stricter consensus reduced errors not simply by assigning fewer cases, but also by preferentially excluding error-enriched cases.

**Fig. 5.**
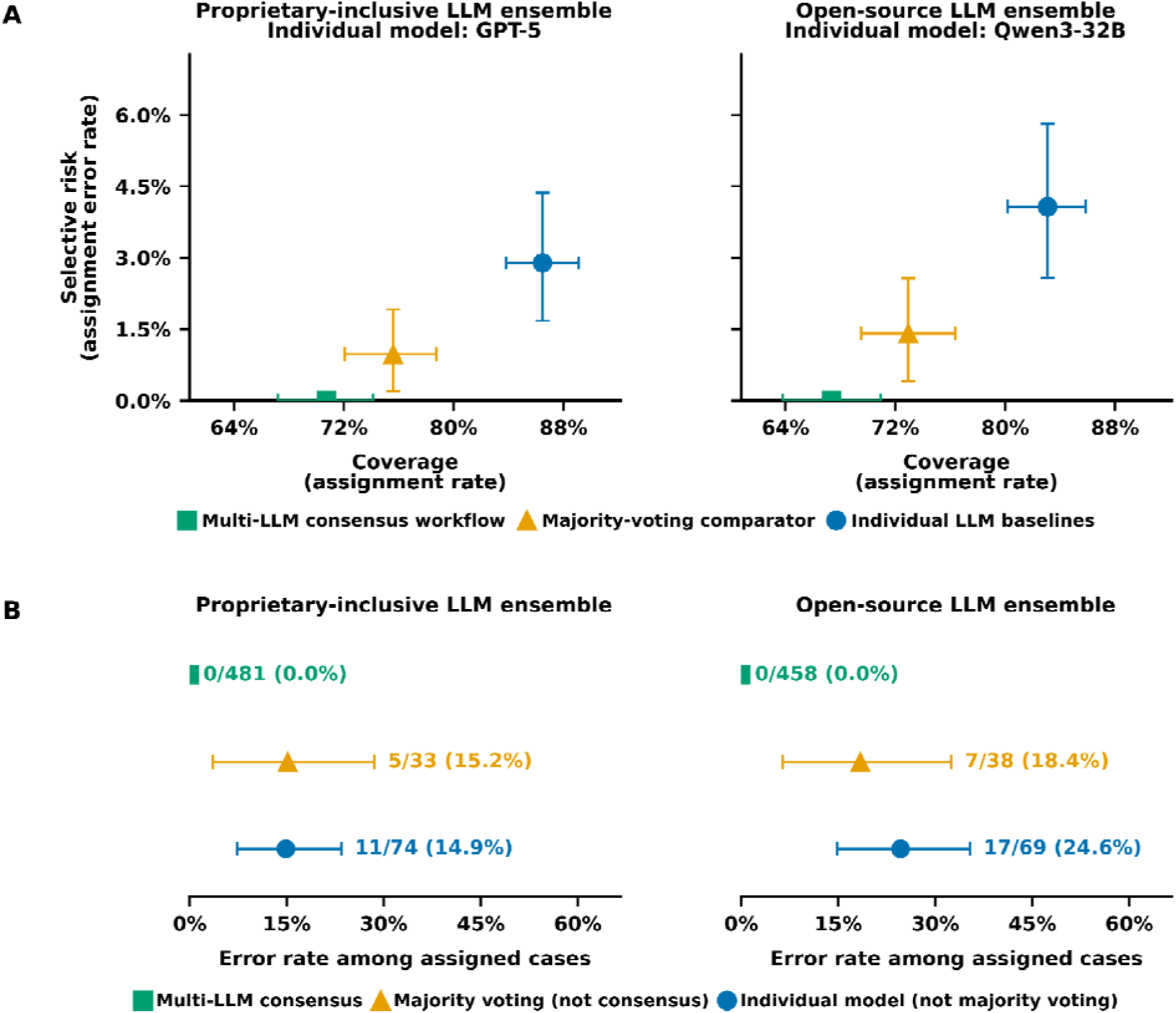
Coverage–risk trade-offs and error enrichment across automatic-assignment rules. Coverage was defined as the proportion of reports automatically assigned as normal or abnormal, and selective risk as the error rate among assigned reports. (A) Coverage and selective risk of the individual-model, majority-voting, and multi-LLM consensus approaches for the proprietary-inclusive (left) and open-source (right) model sets. (B) Error rates were compared among reports assigned under multi-LLM consensus, additional reports assigned after relaxing the rule to majority voting, and further reports assigned after relaxing it to the individual-model baseline. Errors were evaluated using predictions from the corresponding rule. Points represent report-level estimates; error bars indicate 95% confidence intervals from 2,000 patient-level bootstrap replicates. Multi-LLM consensus reduced coverage but yielded no assignment errors in the reference set.

### Within-institution ICD-10 consistency analysis

To compare clinical diagnosis patterns across the report-derived groups, we measured the prevalence of active CNS-related ICD-10 diagnoses in the normal, uncertain, and abnormal groups in the full cohort (Fig. 6). In the proprietary-inclusive ensemble, prevalence increased from 1.6% in the normal group to 29.7% in the uncertain group and 51.3% in the abnormal group. In the open-source ensemble, prevalence was 1.6% in the normal group, 31.1% in the uncertain group, and 51.6% in the abnormal group. In both ensembles, the uncertain group showed an intermediate prevalence between the normal and abnormal groups.

**Fig. 6.**
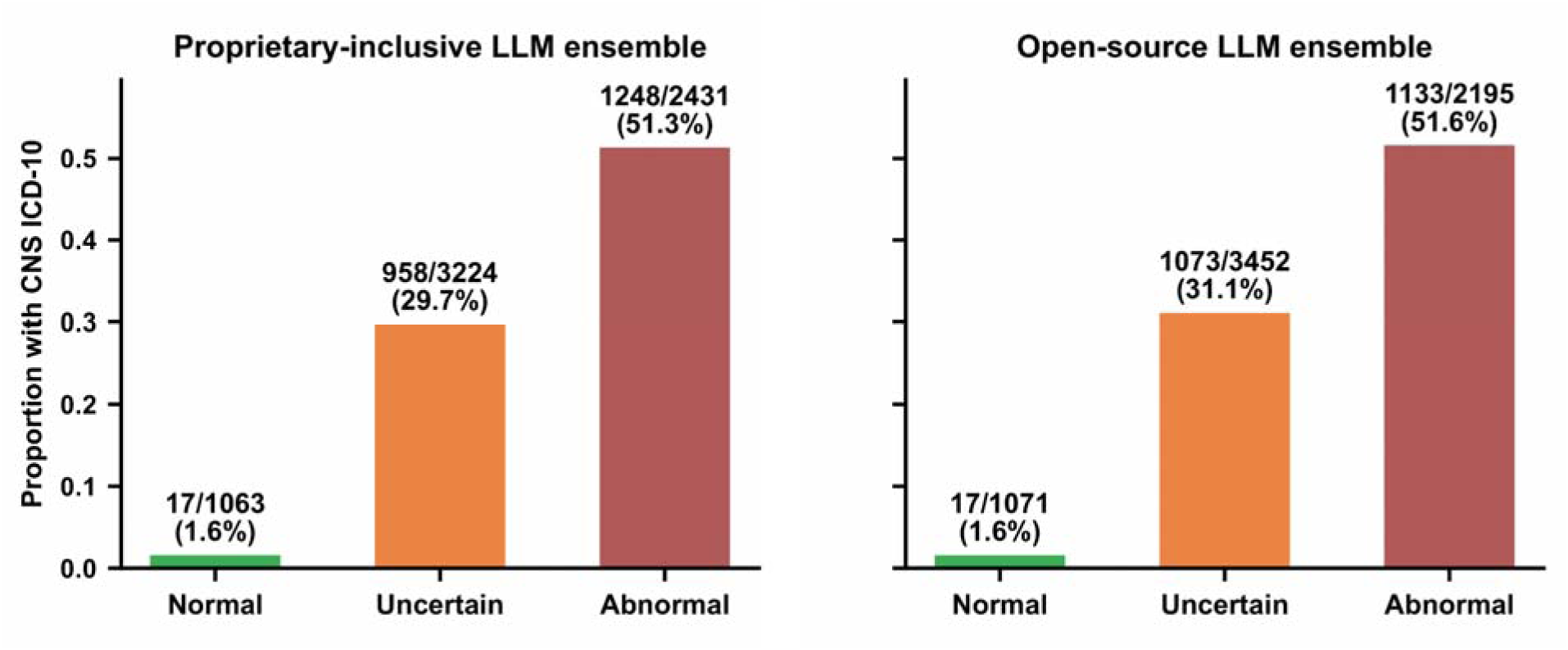
Prevalence of active CNS-related ICD-10 diagnoses across multi-LLM consensus output groups. Bars show the proportion of reports with active CNS-related ICD-10 diagnoses in the assigned normal, uncertain, and assigned abnormal groups of the full cohort for the proprietary-inclusive (left) and open-source (right) ensembles. Labels indicate n/N (%). Prevalence increased stepwise from normal to uncertain to abnormal for both ensembles.

### Review of assigned normal reports with active CNS-related ICD-10 diagnoses

We manually reviewed the 17 reports assigned as normal despite having an active CNS-related ICD-10 diagnosis; the same 17 cases were identified by both ensembles (Additional file 1: Fig. S2). In most cases, the discordance reflected a clinical suspicion or rule-out indication without a corresponding abnormality on the index MRI (n = 13). Other explanations included a prior neurologic condition without a current imaging correlate (n = 2) and CNS pathology outside the brain or not visible on the index MRI (n = 2). In these discordant cases, active CNS-related ICD-10 diagnoses generally reflected clinical or coding context rather than intracranial abnormalities missed by the workflow.

## Discussion

This study evaluated a multi-LLM consensus workflow designed to enable selective semi-automated classification of brain MRI examinations while avoiding forced assignment of ambiguous cases. By requiring unanimous normal or abnormal agreement and classifying WMH-only or model-discordant reports as uncertain, the workflow automatically assigned approximately half of the full cohort while achieving 100% assignment accuracy on the reference set. Compared with individual models, multi-LLM consensus improved abnormal precision, eliminated incorrect assignments, and deferred all expert WMH-only reports in the reference set. These findings suggest that selective consensus can improve assignment reliability while deferring ambiguous or error-prone examinations for manual review.

Consensus outputs may support more precise examination-level cohort construction than ICD-10 screening alone. Assigned normal labels reduce contamination by visible intracranial abnormalities, whereas assigned abnormal labels avoid forced inclusion of WMH-only or ambiguous reports. The uncertain group provides a targeted set for manual review rather than simply representing lost coverage. The stepwise increase in active CNS-related ICD-10 prevalence from the normal to abnormal groups further suggests that the uncertain group represents a clinically mixed review population rather than a nonspecific residual category. This distinction matters because coding variability, missingness, and phenotype misclassification can affect association analyses, case-control comparisons, and imaging biomarker studies [25, 27, 60, 61]. ICD-10 codes remain useful for clinical characterization and cohort-level comparison, whereas report-derived classifications more directly represent findings from a specific brain MRI examination [19, 20, 28, 62].

Building on previous studies demonstrating the feasibility of LLM-based radiology report classification [18–20, 29, 45–47], our findings show that multi-LLM consensus can be used not only to assign labels but also to identify cases requiring manual review. Individual LLMs and majority voting remained more vulnerable to incorrect assignments, whereas requiring unanimous agreement reduced coverage in favor of greater assignment reliability. By treating model disagreement as informative rather than forcing a majority decision, the consensus workflow preferentially deferred cases for which classification was less reliable [63, 64], consistent with selective prediction and clinician-in-the-loop frameworks that reserve uncertain cases for review [65–69]. The two deferral signals played complementary roles. Explicit WMH-only predictions primarily identified the prespecified ambiguous WMH-only examinations, whereas model disagreement captured additional WMH-only cases as well as a broader mixture of expert-normal and expert-abnormal examinations. Together, these findings suggest that explicit ambiguity recognition and model disagreement provide complementary safeguards against incorrect assignment.

These findings are particularly relevant to the research use of routine clinical imaging archives, which provide large-scale real-world brain MRI data but generally lack research-oriented examination labels [5, 6]. The present workflow addresses one barrier to their use by identifying examinations with concordant radiographically normal or abnormal classifications across multiple LLMs while directing ambiguous cases to manual review. Importantly, an examination classified as radiographically normal should not be equated with a healthy research participant. Clinical history, imaging indication, image quality, acquisition protocol, and study-specific eligibility criteria may still be required before such examinations can be included in a particular research dataset or control cohort [70]. The workflow should therefore be viewed as an initial classification step that can support, but does not by itself complete, downstream cohort construction.

The proprietary-inclusive ensemble achieved a slightly higher assignment rate, whereas both ensembles achieved 100% assignment accuracy, 100% normal and abnormal precision, and 100% WMH-only deferral on the reference set. The open-source ensemble therefore provided a more conservative, fully locally deployable alternative (Table 2). Local deployment enables version pinning, model adaptation, and greater auditability [45, 71–73], with potential advantages for privacy, transparency, governance, reproducibility, and local control [74–77].

This study has several limitations. First, its retrospective, single-institution design limits generalizability across institutions, report templates, MRI protocols, scanners, reporting styles, and populations. External validation in independent health systems is required [78–80]. Second, the 680-report expert-adjudicated reference set was purposefully assembled and enriched for normal reports; it was therefore not representative of the full cohort and had a lower prevalence of active CNS-related ICD-10 diagnoses. The reference set may not capture broader disease complexity, rare abnormalities, or reporting heterogeneity. In addition, selection of the 300 supplemental reference reports was guided by lesion categories derived from DeepSeek-R1-32B outputs, which may have influenced the case mix. Final reference labels, however, were assigned independently by expert reviewers blinded to LLM outputs. Third, labels were derived from radiology reports rather than directly from images; therefore, findings that were not documented or emphasized in the reports could not be captured [19, 20, 31, 81, 82]. Fourth, the coarse examination-level abnormal label encompassed heterogeneous findings and did not distinguish etiology, burden, severity, anatomical distribution, or longitudinal change. Finally, the evaluated models, prompts, and deployment settings represent a fixed implementation of the framework. Because LLM outputs may vary with model version, access mode, input formatting, and local reporting conventions, future studies should evaluate the framework across institutions, model versions, MRI protocols, and prospective review workflows [83–85].

## Conclusions

The multi-LLM consensus workflow enabled high-precision identification of radiographically normal and abnormal brain MRI examinations while deferring WMH-only and model-discordant cases for manual review. By prioritizing assignment reliability over complete automation, this approach may provide a scalable first step toward downstream research cohort construction. Within-institution ICD-10 patterns and manual review of ICD-10-discordant cases supported the clinical coherence of the resulting groups. External validation across institutions, reporting styles, MRI protocols, and prospective human-in-the-loop workflows remains necessary.

## List of abbreviations

bfloat16: Brain floating-point 16-bit format
CNS: Central nervous system
CSF: Cerebrospinal fluid
EHR: Electronic health record
ICD-10: International Classification of Diseases, Tenth Revision
JSON: JavaScript Object Notation
LLM: Large language model
MRI: Magnetic resonance imaging
NLP: Natural language processing
WMH: White matter hyperintensity

## Declarations

### Ethics approval and consent to participate

This retrospective study was approved by the Institutional Review Board of NYU Grossman School of Medicine (IRB nos. i14-01224 and i25-01143). The requirement for informed consent was waived by the Institutional Review Board because the study involved retrospective analysis of existing clinical data.

### Consent for publication

Not applicable.

### Availability of data and materials

Due to patient privacy restrictions and institutional data-use requirements, the electronic health record data analyzed during the current study are not publicly available and cannot be shared directly by the authors. The code used for the workflow and analyses is publicly available at https://github.com/WushuangRui/mri-report-llm-classification.

### Authors’ contributions

WR and JM contributed to data processing, experimentation, analysis, and drafting the manuscript. VS contributed to study design, clinical expertise, and expert review. AB contributed substantial revisions to the manuscript. ML and RRP contributed clinical expertise and expert review. JC, DSN, and EF contributed technical and methodological guidance and interpretation of the results. EF and YS conceived the study and supervised the work. All authors read and approved the final manuscript.

## Acknowledgements

The computational requirements for this work were supported in part by the resources and personnel of the NYU Langone High Performance Computing (HPC) Core.

## Additional file

Additional file 1

File name: Additional_file_1.docx

File format: Microsoft Word (.docx)

Title: Supplementary materials

Description: Contains Supplementary Methods, Supplementary Fig. S1–S2, and Supplementary Tables S1–S8.

