## Supplementary Methods, Figures S1-S2, and Tables S1-S8 for "Large language model consensus for reliable research cohort construction from radiology reports: a retrospective cohort study"

### Supplementary Methods. Prompt used for LLM-based JSON structuring of brain MRI reports

You are a specialized medical data extraction assistant focused on analyzing brain MRI reports. Your task is to extract relevant clinical features from raw medical reports about the brain and present them in a structured JSON format. Follow these guidelines:

1. Always return the extracted information in a valid JSON format.

2. Specify the key and the corresponding data type for each piece of information.

3. Use `null` as the value if any information is missing or not present in the report, but maintain the JSON structure.

4. Extract only information explicitly mentioned in the report without adding assumptions or interpretations.

5. Focus on extracting abnormal findings and clinically relevant information.

6. Be precise in describing locations, using anatomical terms as presented in the report.

7. Do not include duplicate information.

Organize the output using the following keys, descriptions, and data types:

*	**`quality_dec`** (boolean): Indicates whether the report mentions image quality degradation due to patient movement or other reasons. Allowed values: `true`, `false`.

*	**`clinical_indication`** (array of objects): An array containing objects providing basic patient information, medical history, and symptoms mentioned in the report. Each `clinical_indication` object should include:

*	`age` (integer or null): The patient's age as a positive integer (e.g., 74, 43).

*	`gender` (string or null): The patient's gender. Allowed values: `"male"`, `"female"`.

*	`symptom` (array of strings or null): Clinical symptoms of the patient (e.g., `["hemibody dysesthesias"]`, `["recurrent vertigo", "headaches"]`).

*	`history` (array of strings or null): The patient's medical history (e.g., `["left hemisphere stroke"]`, `["demyelinating disease"]`).

*	**`comparison`** (array of objects): Information about previous medical examinations for comparison. Each `comparison` object should include:

*	`date_mri` (string or null): The date of the MRI examination, usually found at the end of the report (e.g., `"6/15/2010"`).

*	`date_pre` (string or null): The date of the previous examination used for comparison (e.g., `"6/15/2010"`).

*	`modality` (string or null): The modality of the previous examination (e.g., `"Neck CT"`, `"Brain MRI"`, `"outside study"`).

*	**`lesions`** (array of objects): An array containing objects that describe each lesion found in the report. Each `lesion` object should include:

*	`type` (string): The type of lesion (e.g., `"Hemorrhage"`, `"arteriovenous malformation"`).

*	`location` (array of dictionaries): The location of the lesion, including side and specific brain regions. Each dictionary inside should include:

*	`side` (string or null): The side of the lesion. Allowed values: `"left"`, `"right"`, `"bilateral"`, `null` (if not specified).

*	`region` (array of strings): Specific brain regions where the lesion is located (e.g., `[{"side": "right", "region": ["medulla", "upper cord"]}, {"side": null, "region": ["ventricles", "sulci", "cisterns"]}]`).

*	`size` (dictionary or null): The size of the lesion, including dimensions and units if provided (e.g., `{"AP": 11, "RL": 18, "SI": 15, "unit": "mm"}`, `{"dimension": "5", "unit": "mm"}`).

*	`radiological_changes` (dictionary): Contains a one-to-one correspondence between image modality and image change information. The key (string) is the modality (allowed values: `"T1"`, `"T2"`, `"FLAIR"`, `"T2/FLAIR"`, `"DWI"`, `"ADC"`, `null` if not mentioned). The value (array of strings) is the corresponding image change (e.g., `{"T2/FLAIR": ["hyperintensity"], "T1": ["hypointensity"]}`, `{"null": ["volume loss"]}`).

*	`characteristics` (array of strings or null): Notable descriptors of the lesion, including those suggesting benignity (e.g., "mild," "benign," "incidental," "age-related," "clinically insignificant") or malignancy (e.g., "mass-like," "infiltrative," "aggressive," "malignant," "suspicious"). Always include "non-specific" if mentioned. Other relevant or noteworthy features should also be listed.

*	`compare_pre` (dictionary or null): Contains a one-to-one correspondence between the comparison result and the degree description information. The key (string) is the comparison result (e.g., `"size of the lesion increase"`, `"hyperintense lesion diminish"`). The value (array) is the corresponding degree description (e.g., `{"size of the lesion increase": ["mild"]}`).

*	`diagnosis` (dictionary or null): Contains a one-to-one correspondence between a diagnosis and its confidence level. The key (string) is the diagnosis (e.g., `"developmental venous anomaly"`, `"demyelinating disease"`). The value (array) is the corresponding confidence level (e.g., `{"occult vascular malformations": ["likely"]}`).

*	If there are multiple lesion objects, check whether there are lesions corresponding to the same pathological process and classify them into the overall process object. For example:

```

{

"process": {

"multiple sclerosis": [

{

"type": "active lesion",

"location": {

"side": "right",

"region": [

"periventricular white matter"

]

},

"radiological_changes": {

"T2/FLAIR": ["hyperintense"],

"T1": ["hypointense"]

}

},

{

"type": "chronic lesion",

"location": {

"side": "left",

"region": [

"corpus callosum"

]

},

"radiological_changes": {

"T2/FLAIR": ["hyperintense"],

"T1": ["hypointense"]

}

}

]

}

}

```

*	**`additional_findings`** (array of objects or null): An array containing objects that describe any other relevant abnormal findings. Each object should include:

*	`finding` (string): The type of finding (e.g., `"mucosal thickening"`, `"atrophy"`).

*	`location` (string): The location of the finding.

*	`benign` (boolean): Judge whether it is more likely to be benign or malignant based on the description. Allowed values: `true`, `false`.

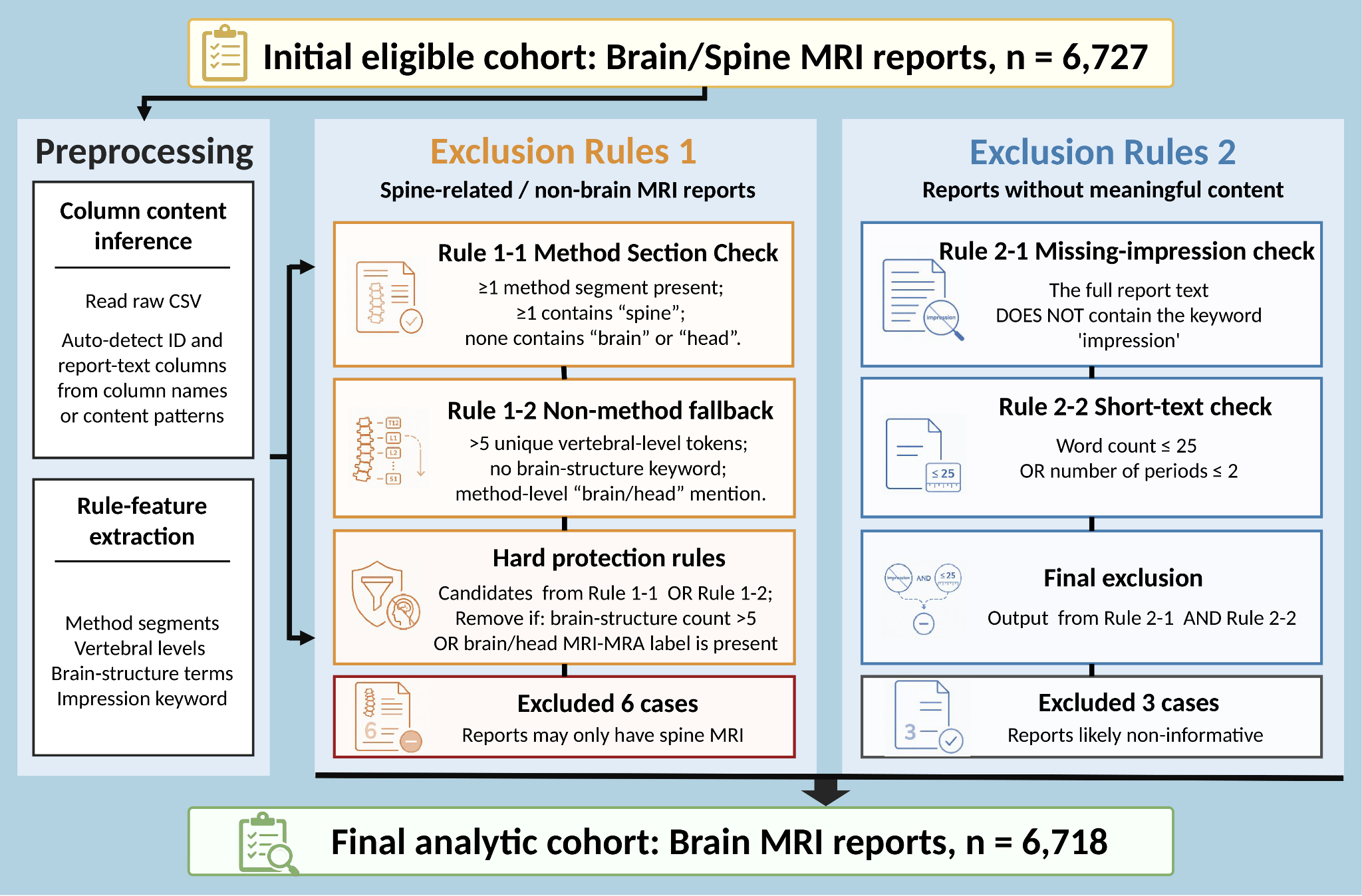

### Supplementary Fig. S1 Rule-based construction of the brain MRI report cohort.

The initial cohort comprised 6,727 brain or spine MRI reports. Preprocessing identified report columns and extracted features used by two exclusion-rule sets. The first removed six spine-related or non-brain reports after applying protection rules for brain MRI content. The second removed three reports lacking meaningful diagnostic content based on missing-impression and short-text criteria. The final analytic cohort comprised 6,718 brain MRI reports.

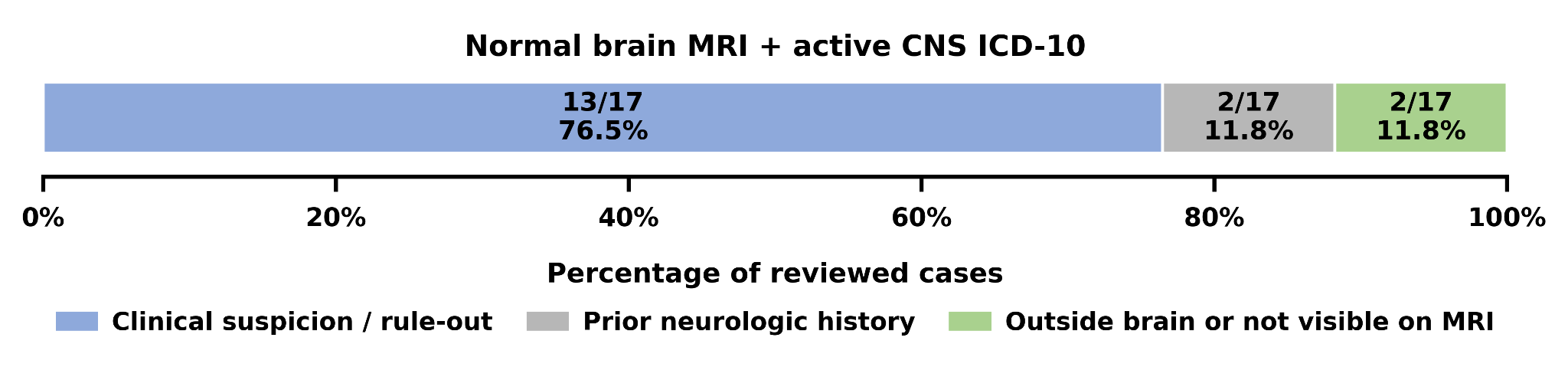

### Supplementary Fig. S2 Explanatory categories for CNS ICD-10 discordance among assigned Normal reports.

The stacked bar shows the distribution of explanations identified during manual review of 17 assigned normal brain MRI reports with CNS-related ICD-10 diagnoses. Discordances were attributed to clinical suspicion or rule-out indications, prior neurologic history without a current MRI correlate, or CNS pathology outside the brain or not visible on the index examination.

Manual review was performed for the 17 consensus-normal brain MRI reports with CNS-related ICD-10 codes identified by the proprietary-inclusive ensemble; the 17 corresponding discordant cases from the open-source ensemble were the same. Although these codes were recorded as active diagnoses in the EHR, manual review indicated that they could reflect a diagnostic workup or prior clinical condition without a corresponding abnormality on the index MRI. As shown in **Fig. S2**, the largest category was **clinical suspicion/rule-out**, which included 13 cases in which the ICD-10 code reflected the clinical indication or diagnostic concern, such as suspected stroke, transient ischemic attack (TIA), dizziness, headache, or demyelinating disease workup, but no corresponding abnormality was present on the index brain MRI. **Prior neurologic history** accounted for 2 cases, in which a historical neurologic diagnosis or previously documented intracranial abnormality had no active or residual imaging correlate on the reviewed MRI. The remaining 2 cases were classified as **outside brain or not visible on MRI**, including conditions involving the spinal cord or optic nerve, where a CNS-related ICD-10 diagnosis could be clinically valid despite the absence of brain parenchymal abnormalities on the index MRI. These findings indicate that ICD-10 discordance primarily reflected the limitations of diagnosis codes as scan-level imaging comparators rather than systematically missed intracranial disease.

### Supplementary Table S1. Definitions and ICD-10 code mappings for the seven CNS-related diagnosis categories used in cohort characterization

| **Category** | **Definition** | **Included ICD-10 codes** | **Representative conditions** |
| --- | --- | --- | --- |
| Vascular | Cerebrovascular disorders involving ischemic or hemorrhagic injury | F01; G08; G45; G46; I60; I61; I62; I63; I65; I66; I67; I68; I69; P52 | Ischemic stroke, intracranial hemorrhage |
| Traumatic | Traumatic intracranial injuries | P10; S02; S04; S06; S09; S12; S14; S24; S34 | Cerebral contusion, subdural hematoma |
| Neoplastic | Primary or secondary intracranial neoplasms | C70; C71; C72; C79.31; D32; D33; D33.3; D42; D43 | Glioma, meningioma, brain metastasis |
| Infectious/inflammatory/immune-mediated | CNS infections, inflammatory disorders, and immune-mediated or demyelinating diseases | A17; A80; A81; A81.2; A83; A86; A87; A88; A89; B00.4; B91; G00; G01; G02; G03; G04; G05; G06; G06.0; G09; G14; G35; G36; G37; G61; G65; G70 | Encephalitis, abscess, multiple sclerosis |
| Congenital/developmental | Congenital malformations and developmental disorders affecting the CNS | G80; Q01; Q02; Q03; Q04; Q04.0; Q05; Q06; Q07; Q85 | Congenital brain malformations, developmental disorders |
| CSF circulation/structural | Disorders involving cerebrospinal fluid circulation or other intracranial structural abnormalities | G91; G93; G94; G95; G96; G97; P90; P91; R90 | Hydrocephalus, structural malformation |
| Neurodegenerative | Degenerative disorders affecting the brain or central nervous system | F02; F03; G10; G11; G12; G13; G20; G21; G23; G30; G31; G32 | Alzheimer disease, other neurodegenerative disorders |

ICD-10 codes were grouped into seven CNS-related diagnostic categories for cohort characterization. Representative conditions are illustrative and not exhaustive.

### Supplementary Table S2. Ranked lesion descriptors used for reference-set enrichment

| **Selection rank** | **Extracted lesion descriptor** | **Count** |  | **Selection rank** | **Extracted lesion descriptor** | **Count** |  | **Selection rank** | **Extracted lesion descriptor** | **Count** |
| --- | --- | --- | --- | --- | --- | --- | --- | --- | --- | --- |
| 1 | demyelinating lesion | 312 |  | 26 | white matter hyperintensities | 49 |  | 51 | chronic small vessel ischemic disease | 26 |
| 2 | white matter lesion | 286 |  | 27 | gliosis | 49 |  | 52 | cyst | 25 |
| 3 | microvascular ischemic disease | 268 |  | 28 | cavernous malformation | 47 |  | 53 | chronic lacunar infarct | 25 |
| 4 | lesion | 234 |  | 29 | hyperintense foci | 43 |  | 54 | white matter disease | 25 |
| 5 | developmental venous anomaly | 158 |  | 30 | chronic ischemic changes | 42 |  | 55 | demyelinating lesions | 24 |
| 6 | white matter lesions | 138 |  | 31 | atrophy | 41 |  | 56 | white matter changes | 23 |
| 7 | infarct | 136 |  | 32 | cavernoma | 39 |  | 57 | capillary telangiectasia | 23 |
| 8 | hyperintensity | 134 |  | 33 | volume loss | 38 |  | 58 | old infarct | 23 |
| 9 | meningioma | 132 |  | 34 | metastatic lesion | 38 |  | 59 | subacute infarct | 22 |
| 10 | chronic microvascular ischemic disease | 129 |  | 35 | arteriovenous malformation | 37 |  | 60 | chronic lacunar infarcts | 22 |
| 11 | mass | 108 |  | 36 | FLAIR hyperintensity | 35 |  | 61 | chronic microhemorrhage | 22 |
| 12 | microvascular disease | 104 |  | 37 | demyelinating plaque | 34 |  | 62 | nonspecific focus | 21 |
| 13 | aneurysm | 88 |  | 38 | infarction | 34 |  | 63 | demyelinating disease | 21 |
| 14 | microhemorrhage | 86 |  | 39 | tumor | 34 |  | 64 | chronic microvascular ischemic changes | 21 |
| 15 | chronic microvascular disease | 81 |  | 40 | demyelinating plaques | 33 |  | 65 | cystic lesion | 20 |
| 16 | microvascular ischemic changes | 80 |  | 41 | punctate focus | 30 |  | 66 | susceptibility artifact | 19 |
| 17 | enhancing lesion | 74 |  | 42 | hyperintensities | 30 |  | 67 | subdural hematoma | 19 |
| 18 | encephalomalacia | 73 |  | 43 | pineal cyst | 29 |  | 68 | postoperative changes | 19 |
| 19 | acute infarct | 72 |  | 44 | signal abnormality | 29 |  | 69 | new lesion | 18 |
| 20 | punctate foci | 67 |  | 45 | multiple sclerosis | 29 |  | 70 | postsurgical changes | 18 |
| 21 | arachnoid cyst | 64 |  | 46 | acute infarction | 28 |  | 71 | abnormal signal | 17 |
| 22 | nonspecific foci | 64 |  | 47 | stenosis | 27 |  | 72 | nonenhancing lesion | 17 |
| 23 | chronic infarct | 62 |  | 48 | lacunar infarcts | 27 |  | 73 | chronic infarction | 17 |
| 24 | multiple sclerosis lesion | 54 |  | 49 | microhemorrhages | 27 |  | 74 | white matter signal abnormality | 17 |
| 25 | lacunar infarct | 51 |  | 50 | hemorrhage | 26 |  | 75 | metastasis | 17 |

| **Selection rank** | **Extracted lesion descriptor** | **Count** |  | **Selection rank** | **Extracted lesion descriptor** | **Count** | |  | | **Selection rank** | **Extracted lesion descriptor** | **Count** |
| --- | --- | --- | --- | --- | --- | --- | --- | --- | --- | --- | --- | --- |
| 76 | punctate hyperintensities | 16 |  | 101 | enhancing mass | 12 |  | | 126 | | microangiopathy | 9 |
| 77 | hyperintense lesion | 16 |  | 102 | glioblastoma | 12 |  | | 127 | | FLAIR signal abnormality | 9 |
| 78 | hypertensive microvascular disease | 16 |  | 103 | ischemic changes | 11 |  | | 128 | | T2/FLAIR hyperintensity | 8 |
| 79 | cord lesion | 16 |  | 104 | vestibular schwannoma | 11 |  | | 129 | | T2 hyperintense lesion | 8 |
| 80 | infundibulum | 15 |  | 105 | lipoma | 11 |  | | 130 | | ischemia | 8 |
| 81 | chronic microangiopathic changes | 14 |  | 106 | saccular aneurysm | 11 |  | | 131 | | restricted diffusion | 8 |
| 82 | microangiopathic changes | 14 |  | 107 | T2/FLAIR signal abnormality | 11 |  | | 132 | | Chiari I malformation | 8 |
| 83 | mesial temporal sclerosis | 14 |  | 108 | focus | 11 |  | | 133 | | microvascular changes | 8 |
| 84 | chronic lesion | 14 |  | 109 | white matter hyperintensity | 11 |  | | 134 | | communicating hydrocephalus | 8 |
| 85 | gliosis/encephalomalacia | 14 |  | 110 | hemorrhagic lesion | 11 |  | | 135 | | chronic lacunar infarction | 8 |
| 86 | hydrocephalus | 14 |  | 111 | cystic encephalomalacia | 10 |  | | 136 | | microvascular ischemic change | 8 |
| 87 | lacunar infarction | 14 |  | 112 | demyelination | 10 |  | | 137 | | remote infarct | 8 |
| 88 | hyperintense signal | 14 |  | 113 | T2 hyperintensity | 10 |  | | 138 | | prominent perivascular spaces | 8 |
| 89 | hematoma | 13 |  | 114 | encephalomalacia gliosis | 10 |  | | 139 | | small foci | 8 |
| 90 | subarachnoid hemorrhage | 13 |  | 115 | occlusion | 10 |  | | 140 | | necrotic mass | 8 |
| 91 | susceptibility | 13 |  | 116 | dilated perivascular spaces | 10 |  | | 141 | | glioma | 8 |
| 92 | ventriculomegaly | 13 |  | 117 | gray matter heterotopia | 10 |  | | 142 | | white matter microangiopathy | 8 |
| 93 | focal cortical dysplasia | 13 |  | 118 | hemorrhagic mass | 10 |  | | 143 | | leptomeningeal enhancement | 8 |
| 94 | chronic microhemorrhages | 12 |  | 119 | T2/FLAIR hyperintense foci | 9 |  | | 144 | | hemosiderin deposition | 7 |
| 95 | mass lesion | 12 |  | 120 | enhancement | 9 |  | | 145 | | venous angioma | 7 |
| 96 | chronic small vessel ischemia | 12 |  | 121 | T2 hyperintensities | 9 |  | | 146 | | narrowing | 7 |
| 97 | parenchymal volume loss | 12 |  | 122 | T2/FLAIR hyperintense lesion | 9 |  | | 147 | | foci | 7 |
| 98 | encephalomalacia/gliosis | 12 |  | 123 | chronic white matter changes | 9 |  | | 148 | | chronic hemorrhage | 7 |
| 99 | white matter injury | 12 |  | 124 | outpouching | 9 |  | | 149 | | susceptibility foci | 7 |
| 100 | subdural collection | 12 |  | 125 | chronic ischemic microvascular disease | 9 |  | | 150 | | encephalomalacia and gliosis | 7 |

| **Selection rank** | **Extracted lesion descriptor** | **Count** |  | **Selection rank** | **Extracted lesion descriptor** | **Count** |  | **Selection rank** | **Extracted lesion descriptor** | **Count** |
| --- | --- | --- | --- | --- | --- | --- | --- | --- | --- | --- |
| 151 | old hemorrhage | 7 |  | 176 | epidermoid cyst | 6 |  | 201 | surgical cavity | 5 |
| 152 | FLAIR hyperintensities | 7 |  | 177 | intraparenchymal hemorrhage | 6 |  | 202 | superficial siderosis | 5 |
| 153 | subdural hygroma | 7 |  | 178 | microangiopathic white matter disease | 6 |  | 203 | fluid collection | 5 |
| 154 | necrotic enhancing mass | 7 |  | 179 | enhancing tumor | 6 |  | 204 | scattered foci | 5 |
| 155 | chronic microangiopathy | 7 |  | 180 | posttreatment changes | 6 |  | 205 | cavitation | 5 |
| 156 | satellite lesion | 7 |  | 181 | occult vascular malformations | 5 |  | 206 | polymicrogyria | 5 |
| 157 | intra-axial lesion | 7 |  | 182 | chronic ischemic white matter changes | 5 |  | 207 | hemorrhagic infarct | 5 |
| 158 | extra-axial fluid collection | 7 |  | 183 | old infarcts | 5 |  | 208 | expansile lesion | 5 |
| 159 | dural based mass | 7 |  | 184 | dural thickening | 5 |  | 209 | residual tumor | 5 |
| 160 | nonspecific punctate foci | 6 |  | 185 | dilated perivascular space | 5 |  | 210 | ring-enhancing lesion | 5 |
| 161 | Wallerian degeneration | 6 |  | 186 | subacute infarction | 5 |  | 211 | chronic hemosiderin deposition | 4 |
| 162 | plaque | 6 |  | 187 | chronic infarcts | 5 |  | 212 | chronic white matter foci | 4 |
| 163 | nonspecific lesion | 6 |  | 188 | sebaceous cyst | 5 |  | 213 | active lesion | 4 |
| 164 | punctate hyperintensity | 6 |  | 189 | microadenoma | 5 |  | 214 | T2/FLAIR hyperintensities | 4 |
| 165 | abnormal signal foci | 6 |  | 190 | remote hemorrhage | 5 |  | 215 | chronic microangiopathic change | 4 |
| 166 | subdural hemorrhage | 6 |  | 191 | hypoplasia | 5 |  | 216 | chronic small vessel disease | 4 |
| 167 | intraventricular hemorrhage | 6 |  | 192 | FLAIR hyperintense lesion | 5 |  | 217 | acute lacunar infarct | 4 |
| 168 | punctate lesion | 6 |  | 193 | nonspecific hyperintensity | 5 |  | 218 | hippocampal cysts | 4 |
| 169 | punctate focus of susceptibility | 6 |  | 194 | focus of susceptibility | 5 |  | 219 | hemangioma | 4 |
| 170 | pineal gland cyst | 6 |  | 195 | pituitary adenoma | 5 |  | 220 | chronic plaques | 4 |
| 171 | susceptibility focus | 6 |  | 196 | epidural hematoma | 5 |  | 221 | abnormal signal intensity | 4 |
| 172 | heterogeneously enhancing lesion | 6 |  | 197 | extra-axial collection | 5 |  | 222 | contrast enhancement | 4 |
| 173 | chronic microvascular ischemia | 6 |  | 198 | bright FLAIR signal | 5 |  | 223 | dilatation | 4 |
| 174 | chronic microvascular ischemic angiopathy | 6 |  | 199 | nodule | 5 |  | 224 | optic neuritis | 4 |
| 175 | chronic microvascular ischemic change | 6 |  | 200 | chronic microvascular angiopathy | 5 |  | 225 | white matter signal abnormalities | 4 |

| **Selection rank** | **Extracted lesion descriptor** | **Count** |  | **Selection rank** | **Extracted lesion descriptor** | **Count** |  | **Selection rank** | **Extracted lesion descriptor** | **Count** |
| --- | --- | --- | --- | --- | --- | --- | --- | --- | --- | --- |
| 226 | enhancing focus | 4 |  | 251 | high-grade glioma | 4 |  | 276 | perivascular space | 3 |
| 227 | arachnoid granulation | 4 |  | 252 | low-grade glioma | 4 |  | 277 | acute ischemia | 3 |
| 228 | chronic small vessel ischemic changes | 4 |  | 253 | atherosclerosis | 4 |  | 278 | chronic microangiopathic ischemic change | 3 |
| 229 | thrombosis | 4 |  | 254 | infiltrative mass | 4 |  | 279 | susceptibility blooming | 3 |
| 230 | nonspecific signal abnormality | 4 |  | 255 | microvascular change | 4 |  | 280 | old blood products | 3 |
| 231 | subacute infarcts | 4 |  | 256 | oligodendroglioma | 4 |  | 281 | thrombus | 3 |
| 232 | petechial hemorrhage | 4 |  | 257 | necrotic lesion | 4 |  | 282 | blood products | 3 |
| 233 | chronic encephalomalacia | 4 |  | 258 | prominent perivascular space | 3 |  | 283 | chronic injury | 3 |
| 234 | encephalocele | 4 |  | 259 | chronic communicating hydrocephalus | 3 |  | 284 | small vessel disease | 3 |
| 235 | subcortical lesion | 4 |  | 260 | microvascular ischemia | 3 |  | 285 | syrinx | 3 |
| 236 | dissection | 4 |  | 261 | ventricular enlargement | 3 |  | 286 | cysts | 3 |
| 237 | new lesions | 4 |  | 262 | T2 hyperintense lesions | 3 |  | 287 | laminar necrosis | 3 |
| 238 | focal outpouching | 4 |  | 263 | hyperintense focus | 3 |  | 288 | cord injury | 3 |
| 239 | cystic structure | 4 |  | 264 | possible lesion | 3 |  | 289 | dolichoectasia | 3 |
| 240 | calcification | 4 |  | 265 | subcortical white matter hyperintensities | 3 |  | 290 | remote microhemorrhage | 3 |
| 241 | multiple sclerosis lesions | 4 |  | 266 | T2 hyperintense foci | 3 |  | 291 | infarcts | 3 |
| 242 | colloid cyst | 4 |  | 267 | chronic changes | 3 |  | 292 | subcortical white matter lesion | 3 |
| 243 | abnormal signal intensity lesion | 4 |  | 268 | focal signal abnormality | 3 |  | 293 | axonopathic injury | 3 |
| 244 | Rathke's cleft cyst | 4 |  | 269 | osteoma | 3 |  | 294 | focal lesion | 3 |
| 245 | ischemic infarct | 4 |  | 270 | chronic appearing changes | 3 |  | 295 | FLAIR hyperintense signal lesion | 3 |
| 246 | extra-axial mass | 4 |  | 271 | T2/FLAIR signal hyperintensity | 3 |  | 296 | old stroke | 3 |
| 247 | recurrent enhancing lesion | 4 |  | 272 | intraosseous hemangioma | 3 |  | 297 | ischemic white matter changes | 3 |
| 248 | primary glial neoplasm | 4 |  | 273 | cerebellar tonsillar ectopia | 3 |  | 298 | white matter focus | 3 |
| 249 | glial neoplasm | 4 |  | 274 | nonspecific hyperintensities | 3 |  | 299 | nonenhancing focus | 3 |
| 250 | resection cavity | 4 |  | 275 | lacunes | 3 |  | 300 | sclerosis | 3 |

Extracted lesion descriptors are listed by selection rank for reference-set enrichment; counts indicate descriptor frequency. FLAIR, fluid-attenuated inversion recovery.

### Supplementary Table S3. ICD-10 diagnosis groups used to define CNS-related codes with potential brain MRI correlates.

| **Disease group** | **ICD-10 code prefix** | **ICD-10 diagnosis category** | **Potential brain MRI correlate / note** |
| --- | --- | --- | --- |
| Cerebrovascular disease | I63 | Cerebral infarction | Acute, subacute, or chronic infarct; encephalomalacia; gliosis; diffusion restriction depending on timing. |
| Cerebrovascular disease | I61 | Intracerebral hemorrhage | Intraparenchymal hemorrhage, hemosiderin deposition, encephalomalacia, or other hemorrhagic sequelae. |
| Traumatic brain injury / intracranial hemorrhage | S06.3 | Focal brain injury | Traumatic contusion, hemorrhagic injury, encephalomalacia, gliosis, or other focal post-traumatic brain abnormality. |
| Traumatic brain injury / intracranial hemorrhage | S06.4 | Epidural hemorrhage | Extra-axial epidural blood products or post-hemorrhagic sequelae. |
| Traumatic brain injury / intracranial hemorrhage | S06.5 | Traumatic subdural hemorrhage | Subdural hematoma, chronic subdural collection, or post-traumatic extra-axial hemorrhagic sequelae. |
| Intracranial neoplasm / mass lesion | C71 | Malignant neoplasm of brain | Intra-axial brain mass, infiltrative tumor, edema, enhancement, mass effect, or treatment-related changes. |
| Intracranial neoplasm / mass lesion | D32 | Benign neoplasm of meninges | Meningioma or other dural-based mass; may cause mass effect or adjacent parenchymal changes. |
| Intracranial neoplasm / mass lesion | C79.31 | Secondary malignant neoplasm of brain | Brain metastasis, leptomeningeal disease, edema, enhancement, or treatment-related changes. |
| Intracranial neoplasm / mass lesion | C83.390 | Primary central nervous system lymphoma | Intracranial mass or infiltrative lesion, often with enhancement and restricted diffusion; findings depend on disease status and prior treatment. |
| Intracranial infection / inflammatory disease | G06.0 | Intracranial abscess and granuloma | Ring-enhancing abscess, granulomatous lesion, edema, restricted diffusion, or mass effect. |
| Intracranial infection / inflammatory disease | G04.0 | Acute disseminated encephalitis | Multifocal inflammatory or demyelinating brain lesions; may overlap with ADEM-like imaging patterns. |
| Intracranial infection / inflammatory disease | B00.4 | Herpesviral encephalitis | Temporal lobe or limbic signal abnormality, diffusion restriction, enhancement, or edema. |
| CSF / structural abnormality | G91 | Hydrocephalus | Ventriculomegaly, transependymal CSF flow, aqueductal obstruction, or shunt-related findings. |
| Intracranial infection / inflammatory disease | A81.2 | Progressive multifocal leukoencephalopathy | Multifocal white matter lesions, often non-enhancing or minimally enhancing depending on stage. |
| Demyelinating disease | G35 | Multiple sclerosis | White matter demyelinating lesions, enhancing active lesions, or chronic demyelinating plaques; may be absent on index brain MRI in some clinical contexts. |
| Demyelinating disease | G36 | Other acute disseminated demyelination | Demyelinating lesions may involve brain, optic nerve, or spinal cord; brain MRI can be normal if disease is outside the brain. |
| Cranial nerve / skull base lesion | D33.3 | Benign neoplasm of cranial nerves | Cranial nerve or cerebellopontine angle lesion, such as vestibular schwannoma; may be outside brain parenchyma. |
| Congenital / structural brain abnormality | Q04.0 | Congenital malformations of corpus callosum | Agenesis, dysgenesis, or hypoplasia of the corpus callosum, potentially accompanied by other congenital brain abnormalities. |

The table lists ICD-10 code prefixes selected a priori to identify CNS-related diagnoses that could plausibly have visible correlates on brain MRI. These codes were used as a within-institution clinical comparator rather than as scan-level ground truth.

| Supplementary Table S4. Precision, recall, and assignment performance across classification strategies | | | | | |
| --- | --- | --- | --- | --- | --- |
| **Classification strategy** | **Normal precision** | **Normal recall** | **Abnormal precision** | **Abnormal recall** | **Automatic assignment rate** |
| **ICD-10-based comparator** | | | | | |
| ICD-10 binary classification | 0.658 (0.618–0.697) | 0.995 (0.986–1.000) | 0.941 (0.896–0.982) | 0.509 (0.441–0.577) | 1.000 (1.000–1.000) |
| **Individual LLM baselines** | | | | | |
| Qwen3-32B | 0.948 (0.925–0.969) | 0.987 (0.974–0.997) | 0.983 (0.962–1.000) | 0.803 (0.749–0.854) | 0.831 (0.802–0.860) |
| Kimi-Linear-48B-A3B | 0.985 (0.970–0.997) | 0.871 (0.836–0.903) | 0.844 (0.794–0.891) | 0.844 (0.791–0.893) | 0.804 (0.774–0.834) |
| GPT-5 | 0.989 (0.977–0.997) | 0.978 (0.963–0.992) | 0.941 (0.908–0.970) | 0.950 (0.918–0.977) | 0.865 (0.839–0.891) |
| DeepSeek-R1-32B | 0.936 (0.910–0.959) | 0.981 (0.966–0.994) | 0.964 (0.933–0.988) | 0.734 (0.673–0.792) | 0.818 (0.789–0.848) |
| **Majority-voting comparator** | | | | | |
| Proprietary-inclusive LLM ensemble | 0.997 (0.991–1.000) | 0.922 (0.893–0.948) | 0.976 (0.952–0.994) | 0.761 (0.706–0.817) | 0.756 (0.723–0.789) |
| Open-source LLM ensemble | 0.988 (0.977–0.997) | 0.922 (0.894–0.948) | 0.980 (0.955–1.000) | 0.670 (0.604–0.732) | 0.729 (0.695–0.764) |
| **Multi-LLM consensus workflow** | | | | | |
| Proprietary-inclusive LLM ensemble | 1.000 (1.000–1.000) | 0.858 (0.822–0.891) | 1.000 (1.000–1.000) | 0.743 (0.686–0.800) | 0.707 (0.673–0.743) |
| Open-source LLM ensemble | 1.000 (1.000–1.000) | 0.863 (0.828–0.896) | 1.000 (1.000–1.000) | 0.628 (0.563–0.693) | 0.674 (0.639–0.708) |

Values are point estimates with 95% confidence intervals in parentheses. Precision was calculated among reports assigned each label, whereas recall was calculated among reports with the corresponding expert label and therefore incorporated both misclassification and deferral. Increasing consensus stringency improved Normal and Abnormal precision but reduced recall and assignment rate, demonstrating the trade-off between cohort purity and coverage.

### Supplementary Table S5. Expert reference composition of multi-LLM consensus workflow outputs

| **Proprietary-inclusive LLM ensemble** | | | | |
| --- | --- | --- | --- | --- |
| Expert reference label | Assigned Normal | Uncertain | Assigned Abnormal | Total |
| Normal | 319 (100.0%) | 53 (26.6%) | 0 (0.0%) | 372 |
| WMH-only | 0 (0.0%) | 90 (45.2%) | 0 (0.0%) | 90 |
| Abnormal | 0 (0.0%) | 56 (28.1%) | 162 (100.0%) | 218 |
| Total | 319 | 199 | 162 | 680 |
| **Open-source LLM ensemble** | | | | |
| Expert reference label | Assigned Normal | Uncertain | Assigned Abnormal | Total |
| Normal | 321 (100.0%) | 51 (23.0%) | 0 (0.0%) | 372 |
| WMH-only | 0 (0.0%) | 90 (40.5%) | 0 (0.0%) | 90 |
| Abnormal | 0 (0.0%) | 81 (36.5%) | 137 (100.0%) | 218 |
| Total | 321 | 222 | 137 | 680 |

Values are counts with column percentages in parentheses.

### Supplementary Table S6. Confidence-based selective classification performance of individual LLMs at matched assignment rates

| **Classification strategy** | **Assignment rate** | **Assignment accuracy** | **Normal precision** | **Abnormal precision** | **Assignment errors (n)** | **WMH-only deferral rate** |
| --- | --- | --- | --- | --- | --- | --- |
| **Individual LLM selective baselines using model-reported confidence** | | | | | | |
| Qwen3-32B | 0.674 (0.639–0.708) | 0.967 (0.950–0.983) | 0.953 (0.927–0.975) | 0.994 (0.980–1.000) | 15 (8–23) | 0.856 (0.779–0.925) |
| GPT-5 | 0.707 (0.673–0.743) | 0.985 (0.973–0.996) | 0.993 (0.982–1.000) | 0.974 (0.949–0.995) | 7 (2–13) | 0.944 (0.893–0.988) |

To assess whether the performance gain from multi-model consensus could be explained solely by selective deferral, we constructed model-reported confidence baselines using GPT-5 and Qwen3-32B. For each report, the model provided a predicted class and a model-reported confidence score for that prediction. Confidence was elicited directly from each model on a [0–1] scale using the same classification input. Predictions assigned to the prespecified WMH-only category were deferred, whereas Normal and Abnormal predictions were ranked by confidence. For each model, predictions were retained in descending order of confidence until the number of assigned reports matched that of the corresponding three-model unanimous consensus workflow. Values are point estimates with 95% confidence intervals in parentheses; assignment errors are reported as counts. At matched assignment rates, confidence-selected individual models remained less accurate and produced more errors than multi-LLM consensus, indicating that the consensus advantage was not explained by selective deferral alone.

### Supplementary Table S7. Performance of pairwise LLM consensus ensembles

| **Classification strategy** | **Assignment rate** | **Assignment accuracy** | **Normal precision** | **Abnormal precision** | **Assignment errors (n)** | **WMH-only deferral rate** |
| --- | --- | --- | --- | --- | --- | --- |
| **Pairwise LLM consensus ensembles** | | | | | | |
| Qwen3-32B+Kimi-Linear-48B-A3B | 0.712 (0.678–0.744) | 0.994 (0.986–1.000) | 0.994 (0.984–1.000) | 0.994 (0.980–1.000) | 3 (0–7) | 0.978 (0.943–1.000) |
| Qwen3-32B + GPT-5 | 0.797 (0.765–0.827) | 0.989 (0.980–0.996) | 0.989 (0.978–0.997) | 0.989 (0.971–1.000) | 6 (2–11) | 0.944 (0.895–0.988) |
| Qwen3-32B+DeepSeek-R1-32B | 0.766 (0.734–0.799) | 0.981 (0.968–0.992) | 0.976 (0.958–0.990) | 0.993 (0.979–1.000) | 10 (4–17) | 0.933 (0.879–0.979) |
| Kimi-Linear-48B-A3B + GPT-5 | 0.746 (0.712–0.778) | 0.986 (0.975–0.996) | 1.000 (1.000–1.000) | 0.963 (0.934–0.989) | 7 (2–13) | 0.956 (0.907–0.990) |
| Kimi-Linear-48B-A3B + DeepSeek-R1-32B | 0.687 (0.651–0.718) | 0.994 (0.985–1.000) | 0.997 (0.990–1.000) | 0.986 (0.963–1.000) | 3 (0–7) | 0.978 (0.943–1.000) |
| GPT-5 + DeepSeek-R1-32B | 0.772 (0.741–0.803) | 0.987 (0.975–0.994) | 0.989 (0.977–0.997) | 0.981 (0.958–1.000) | 7 (3–13) | 0.933 (0.875–0.978) |

Two-model consensus generally increased assignment rates relative to the three-model consensus workflows, at the cost of slightly lower assignment accuracy and additional assignment errors.

### Supplementary Table S8. Sensitivity analysis of selective classification performance using raw brain MRI reports as model input

| **Classification strategy** | **Assignment rate** | **Assignment accuracy** | **Normal precision** | **Abnormal precision** | **Assignment errors (n)** | **WMH-only deferral rate** |
| --- | --- | --- | --- | --- | --- | --- |
| **Individual LLM selective baselines using raw radiology report input** | | | | | | |
| Qwen3-32B | 0.850 (0.823–0.878) | 0.972 (0.959–0.985) | 0.989 (0.977–0.997) | 0.943 (0.910–0.972) | 16 (9–24) | 0.889 (0.819–0.947) |
| Kimi-Linear-48B-A3B | 0.791 (0.759–0.823) | 0.965 (0.948–0.979) | 0.965 (0.945–0.982) | 0.964 (0.933–0.988) | 19 (11–28) | 0.822 (0.738–0.895) |
| GPT-5 | 0.875 (0.850–0.900) | 0.973 (0.959–0.985) | 0.992 (0.981–1.000) | 0.942 (0.910–0.972) | 16 (9–24) | 0.844 (0.764–0.916) |
| DeepSeek-R1-32B | 0.834 (0.806–0.863) | 0.954 (0.936–0.970) | 0.975 (0.958–0.990) | 0.915 (0.874–0.951) | 26 (17–36) | 0.900 (0.833–0.957) |
| **Majority-voting comparator** | | | | | | |
| Proprietary-inclusive LLM ensemble | 0.768 (0.736–0.800) | 0.992 (0.984–0.998) | 0.994 (0.986–1.000) | 0.988 (0.968–1.000) | 4 (1–8) | 0.956 (0.908–0.989) |
| Open-source LLM ensemble | 0.760 (0.728–0.793) | 0.990 (0.981–0.998) | 0.994 (0.986–1.000) | 0.981 (0.956–1.000) | 5 (1–10) | 0.956 (0.910–0.990) |
| **Multi-LLM consensus workflow** | | | | | | |
| Proprietary-inclusive LLM ensemble | 0.757 (0.723–0.791) | 0.992 (0.984–0.998) | 0.994 (0.985–1.000) | 0.988 (0.968–1.000) | 4 (1–8) | 0.956 (0.908–0.989) |
| Open-source LLM ensemble | 0.734 (0.699–0.769) | 0.992 (0.984–0.998) | 0.994 (0.985–1.000) | 0.987 (0.966–1.000) | 4 (1–8) | 0.956 (0.910–0.990) |

Models were evaluated using raw brain MRI report text rather than the structured JSON input used in the primary analysis, with the same selective classification rules and ensemble definitions. Values are point estimates with 95% confidence intervals in parentheses; assignment errors are reported as counts. Raw reports outperformed JSON-structured reports in assignment accuracy for individual LLMs, whereas multi-model consensus reversed this pattern. Richer raw-report context may improve individual models but also promote correlated errors when models respond similarly to misleading or ambiguous language. JSON structuring may reduce individual-model performance but increase disagreement on difficult cases, thereby improving accuracy among assigned cases.
